# The statistical analysis of daily data associated with different parameters of the New Coronavirus COVID-19 pandemic in Georgia and their short-term interval prediction in spring 2021

**DOI:** 10.1101/2021.06.16.21259038

**Authors:** Avtandil G. Amiranashvili, Ketevan R. Khazaradze, Nino D. Japaridze

**Affiliations:** Mikheil Nodia Institute of Geophysics of Ivane Javakhishvili Tbilisi State University, Tbilisi, Georgia; Georgian State Teaching University of Physical Education and Sport, Tbilisi, Georgia; Ministry of Internally Displaced Persons from Occupied Territories, Labour, Health and Social Affair of Georgia, Tbilisi, Georgia; Tbilisi State Medical University, Tbilisi, Georgia

**Keywords:** New Coronavirus COVID-19, statistical analysis, short-term prediction

## Abstract

The lockdown introduced in Georgia on November 28, 2020 brought positive results. There are clearly positive tendencies in the spread of COVID-19 to February - first half of March 2021. However, in April-May 2021 there was a significant deterioration in the epidemiological situation.

In this work results of the next statistical analysis of the daily data associated with New Coronavirus COVID-19 infection of confirmed (C), recovered (R), deaths (D) and infection rate (I) cases of the population of Georgia in the period from March 01, 2021 to May 31, 2021 are presented. It also presents the results of the analysis of two-week forecasting of the values of C, D and I. The information was regularly sent to the National Center for Disease Control & Public Health of Georgia and posted on the Facebook page https://www.facebook.com/Avtandil1948/.

The analysis of data is carried out with the use of the standard statistical analysis methods of random events and methods of mathematical statistics for the non-accidental time-series of observations. In particular, the following results were obtained.

Georgia’s ranking in the world for Covid-19 monthly mean values of infection and deaths cases in spring 2021 (per 1 million population) was determined. Among 156 countries with population ≥ 1 million inhabitants in May 2021 Georgia was in the 11 place on new infection cases and in the 14 place on Death.

A comparison between the daily mortality from Covid-19 in Georgia in spring 2021 with the average daily mortality rate in 2015-2019 shows, that the largest share value of D from mean death in 2015-2019 was 25.3 % (22.05.2021), the smallest 1.42 % (15.03.2021).

Data about infection rate of the population of Georgia with Covid-19 according to traffic light system shown, that Georgia in April and May 2021 was in the red zone.

The statistical analysis of the daily and decade data associated with coronavirus COVID-19 pandemic of confirmed, recovered, deaths cases and infection rate of the population of Georgia are carried out. Maximum daily values of investigation parameters are following: C = 2171 (05.05.2021), R = 2038 (17.05.2021), D = 33 (22.05.2021), I = 8.05 % (04.05.2020). Maximum mean decade values of investigation parameters are following: C = 1258 (3 Decade of April 2021), R = 1283 (2 Decade of May 2021), D = 24 (2 Decade of May 2021), I = 6.54 % (1 Decade of May 2021).

It was found that as with September 2020 to February 2021 [8], in spring 2021 the regression equations for the time variability of the daily values of C, R and D have the form of a tenth order polynomial.

Mean values of speed of change of confirmed -V(C), recovered - V(R), deaths - V(D) and infection rate V(I) coronavirus-related cases in different decades of months in the spring 2021 were determined. Maximum mean decade values of investigation parameters are following: V(C) = +37 cases/day (1 Decade of April 2021), V(R) = +36 cases/day (3 Decade of April 2021), V(D) = +0.6 cases/day (3 Decade of April 2021), V(I) = + 0.17 %/ day (2 and 3 decades of April 2021).

Cross-correlations analysis between confirmed COVID-19 cases with recovered and deaths cases shows, that the maximum effect of recovery is observed 9 and 13 days after infection, and deaths - after 12-17 days.

Comparison of real and calculated predictions data of C, D and I in Georgia are carried out. It was found that two-week daily and mean two-week real values of C, D and I practically fall into the 67% - 99.99% confidence interval of these predicted values for the specified time periods.

The comparison of data about C and D in Georgia (GEO) with similar data in Armenia (ARM), Azerbaijan (AZE), Russia (RUS), Turkey (TUR) and in the World (WRL) is also carried out.

## 1. Introduction

Almost a year and a half has passed since the outbreak of the novel coronavirus (COVID-19) in China, recognized on March 11, 2020 as a pandemic due to its rapid spread in the world [1]. Until now, despite the measures taken, the overall level of morbidity and mortality remains quite high. During this time, together with epidemiologists, scientists and specialists from various disciplines from all over the world have joined intensive research on this unprecedented phenomenon (including Georgia [2-8]).

In particular, in our works [6-8] it was noted that specialists in the field of physical and mathematical sciences make an important contribution to research on the spread of the new coronavirus COVID-19. Work on systematization, statistical analysis, forecasting, spatial-temporal modeling of the spread of the new coronavirus is being actively continued at the present time [9-15, etc.].

This work is a continuation of the researches [6-8].

In this work results of a statistical analysis of the daily data associated with New Coronavirus COVID-19 infection of confirmed (C), recovered (R), deaths (D) and infection rate (I) cases of the population of Georgia in the period from March 01, 2021 to May 31, 2021 are presented. It also presents the results of the analysis of two-week forecasting of the values of C, D and I. The information was regularly sent to the National Center for Disease Control & Public Health of Georgia and posted on the Facebook page https://www.facebook.com/Avtandil1948/.

The comparison of data about C and D in Georgia with similar data in Armenia, Azerbaijan, Russia, Turkey and in the world is also carried out.

We used standard methods of statistical analysis of random events and methods of mathematical statistics for non-random time series of observations [6-8, 16, 17].

## 2. Study areas, material and methods

The study area: Georgia. Data of John Hopkins COVID-19 Time Series Historical Data (with US State and County data) [https://www.soothsawyer.com/john-hopkins-time-series-data-with-us-state-and-county-city-detail-historical/; https://data.humdata.org/dataset/total-covid-19-tests-performed-by-country] and https://stopcov.ge about daily values of confirmed, recovered, deaths and infection rate coronavirus-related cases, from March 01, 2021 to May 31, 2021 are used. The work also used data of National Statistics Office of Georgia (Geostat) on the average monthly total mortality in Georgia in October-May 2015-2019 [https://www.geostat.ge/en/]. In the proposed work the analysis of data is carried out with the use of the standard statistical analysis methods of random events and methods of mathematical statistics for the non-accidental time-series of observations [6-8, 16, 17].

The following designations will be used below: Mean – average values; Min – minimal values; Max - maximal values; Range – Max-Min; St Dev - standard deviation; σ_m_ - standard error; C_V_= 100·St Dev/Mean – coefficient of variation, %; R^2^ – coefficient of determination; r – coefficient of linear correlation; CR – coefficient of cross correlation; Lag = 1, 2…30 Day; K_DW_ – Durbin-Watson statistic; Calc – calculated data; Real - measured data; D1, D2, D3 – numbers of the month decades; α - the level of significance; C, R, D - daily values of confirmed, recovered and deaths coronavirus-related cases; V(C), V(R) and V(D) - daily values of speed of change of confirmed, recovered and deaths coronavirus-related cases (cases/day); I - daily values of infection rate (or positive rate) coronavirus-related cases (100· C/number of coronavirus tests performed); V(I) - daily values of speed of change of I; DC – deaths coefficient, % = (100· D/C). Official data on number of coronavirus tests performed are published from December 05, 2020 [https://stopcov.ge].

The rate of new infection of the population with Covid-19 according to traffic light system was determined [https://www.dw.com/en/coronavirus-what-the-eus-new-traffic-light-system-means/a-55265476]

The rate of new infections per 100 000 inhabitants in the previous 14 days and the rate of positive covid-19 tests decide which color is attributed to a given region.

- Green is for regions reporting less than 25 new infections per 100 000 inhabitants, and test positivity is below 4%.
- Orange is for regions reporting less than 50 new infections, and test positivity is over 4% — or the incidence is between 25 and 150 and test positivity is below 4%.
- Red is for regions with more than 50 new infections per 100 000 incidents, and test positivity is over 4% — or the incidence is over 150 per 100 000 in the past 14 day.

The statistical programs Data Fit 7, Mesosaur and Excel 16 were used for calculations.

The curve of trend is equation of the regression of the connection of the investigated parameter with the time at the significant value of the determination coefficient and such values of K_DW_, where the residual values are accidental. If the residual values are not accidental the connection of the investigated parameter with the time we will consider as simply regression.

The calculation of the interval prognostic values of C, D and I taking into account the periodicity in the time-series of observations was carried out using Excel 16 (the calculate methodology was description in [7]). The duration of time series of observations for calculating ten-day and two-week forecasts of the values of C, D and I was 35 - 92 days.

67%…99.99%_Low - 67% 99.99% lower level of confidence interval of prediction values of C, D and I; 67%…99.99%_Upp - 67% 99.99% upper level of confidence interval of prediction values of C, D and I.

In the Table 1 [8] the scale of comparing real data with the predicted ones and assessing the stability of the time series of observations in the forecast period in relation to the pre-predicted one (period for prediction calculating) is presented.

**Table 1.** The statistical characteristics of Covid-19 confirmed cases to 1 million populations in Georgia, neighboring countries (Armenia, Azerbaijan, Russia, Turkey) and World from March 1, 2021 to May 31, 2021 (r_min_ = ± 0.21, α = 0.05).

| Parameter | GEO | ARM | AZE | RUS | TUR | WRL |
| --- | --- | --- | --- | --- | --- | --- |
| Mean | 213 | 186 | 107 | 61 | 328 | 79 |
| Min | 36 | 11 | 9 | 52 | 77 | 36 |
| Max | 582 | 424 | 268 | 78 | 748 | 116 |
| Range | 547 | 413 | 259 | 27 | 671 | 80 |
| Median | 182 | 169 | 85 | 60 | 257 | 80 |
| St Dev | 129 | 119 | 76 | 5 | 212 | 21 |
| $\sigma_m$ | 13.6 | 12.4 | 8.0 | 0.6 | 22.2 | 2.2 |
| $C_v$ (%) | 60.6 | 63.8 | 71.5 | 8.9 | 64.4 | 27.2 |
| Correlation Matrix |  |  |  |  |  |  |
| GEO | 1 | -0.09 | 0.14 | -0.57 | 0.19 | 0.72 |
| ARM | -0.09 | 1 | 0.79 | -0.02 | 0.66 | 0.34 |
| AZE | 0.14 | 0.79 | 1 | -0.31 | 0.91 | 0.56 |
| RUS | -0.57 | -0.02 | -0.31 | 1 | -0.34 | -0.60 |
| TUR | 0.19 | 0.66 | 0.91 | -0.34 | 1 | 0.63 |
| WRL | 0.72 | 0.34 | 0.56 | -0.60 | 0.63 | 1 |

## 3. Results and Discussion

The results in the Fig. 1-23 and Table 1-9 are presented.

**Fig. 1.**
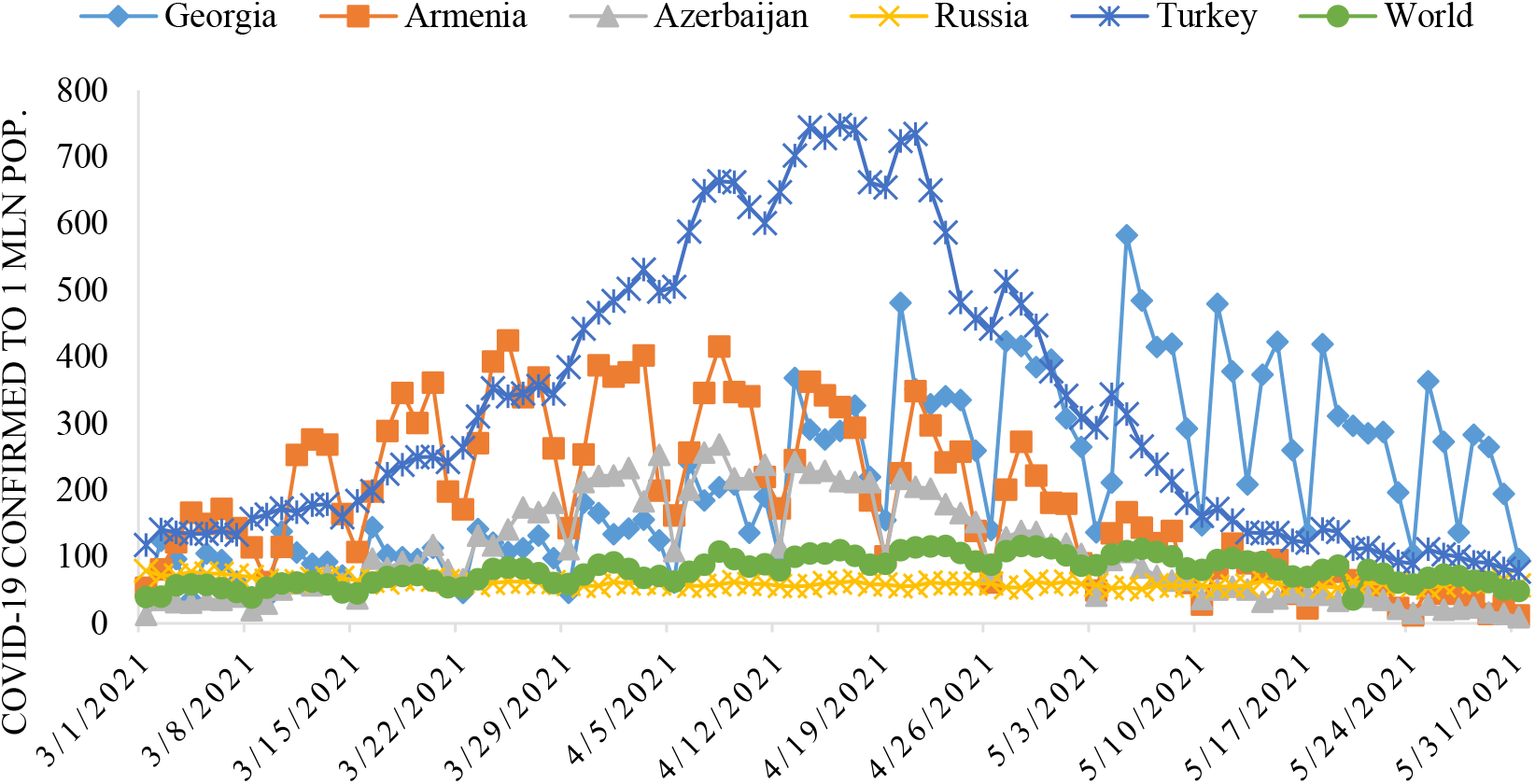
Time-series of Covid-19 confirmed cases to 1 million populations in Georgia, neighboring countries (Armenia, Azerbaijan, Russia, Turkey) and World from March 1, 2021 to May 31, 2021.

### 3.1 Comparison of time-series of Covid-19 confirmed and deaths cases in Georgia, its neighboring countries and World in spring 2021

The time-series curves and statistical characteristics of Covid-19 confirmed and deaths cases (to 1 million populations) in Georgia, its neighboring countries and World from March 01, 2021 to May 31, 2021 in Fig. 1 - 2 and in Table 1-2 are presented.

Variability of the values of C per 1 million populations is as follows (Fig. 1, Table 1):

- Georgia. Range of change: 36-582, mean value - 213.
- Armenia. Range of change: 11-424, mean value - 186.
- Azerbaijan. Range of change: 9-268, mean value - 107.
- Russia. Range of change: 52-78, mean value - 61.
- Turkey. Range of change: 77-748, mean value - 328.
- World. Range of change: 36-116, mean value - 79.

The largest variations in C values were observed in Turkey (C_V_=71.5%), the smallest - in Russia (C_V_=8.9%).

Significant linear correlation (r_min_ = ± 0.21, α = 0.05) between these countries on C value varies from − 0.57 (pair Georgia-Russia) to +0.91 (pair Azerbaijan-Turkey). Linear correlation between World ad these countries is significant and varied from −0.60 (pair World-Russia) to +0.72 (pair World-Georgia). Thus, in addition to Russia, in the rest of these countries, there are more or less similar trends to the world in the development of the coronavirus epidemic (Table 1).

Variability of the values of D per 1 million populations is as follows (Fig. 2, Table 2):

**Table 2.**
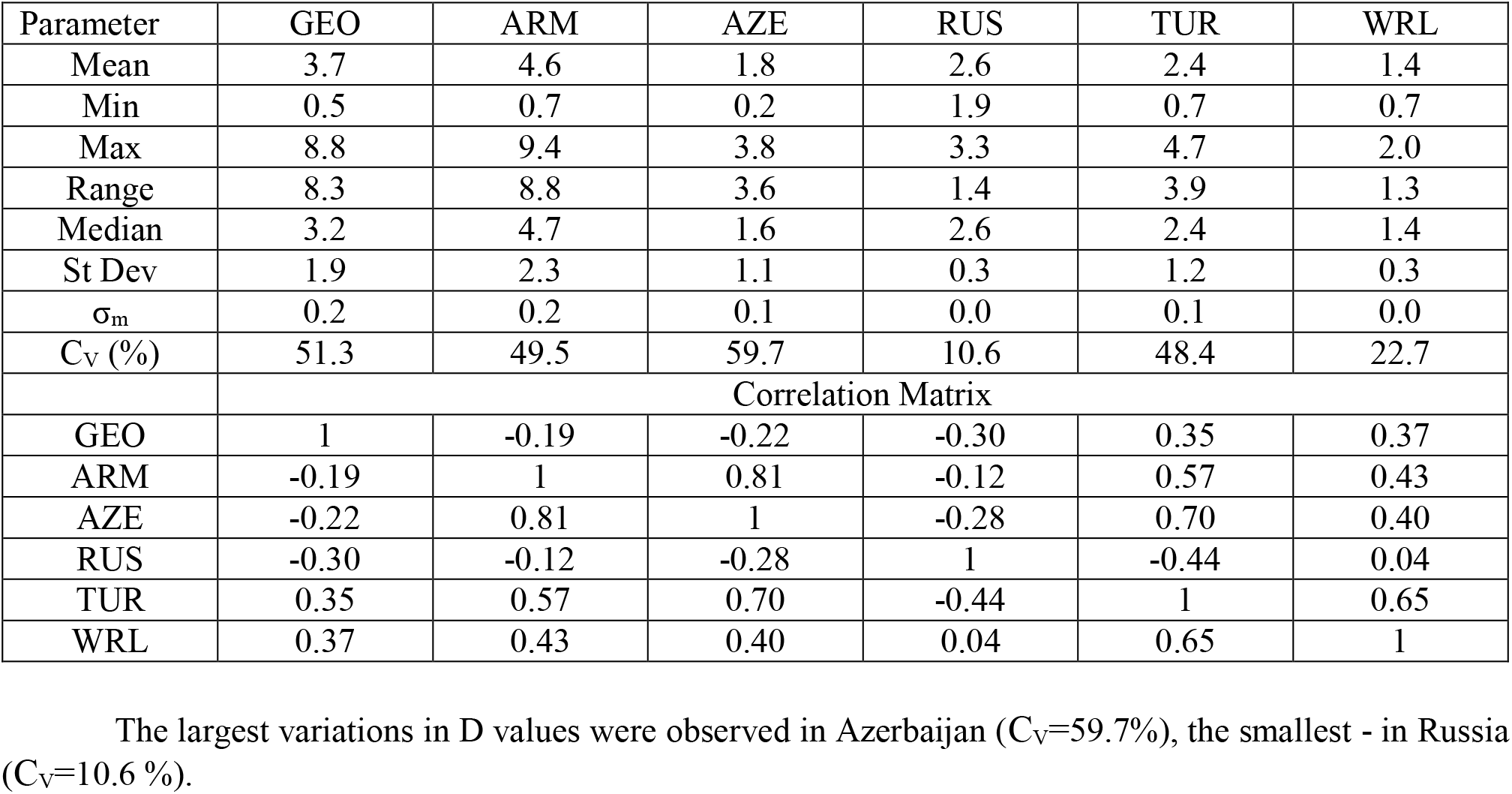
The statistical of deaths cases from Covid-19 to 1 million populations in Georgia, neighboring countries (Armenia, Azerbaijan, Russia, Turkey) and World from March 1, 2021 to May 31, 2021. (r_min_ = ± 0.21, α = 0.05).

| Parameter | GEO | ARM | AZE | RUS | TUR | WRL |
| --- | --- | --- | --- | --- | --- | --- |
| Mean | 3.7 | 4.6 | 1.8 | 2.6 | 2.4 | 1.4 |
| Min | 0.5 | 0.7 | 0.2 | 1.9 | 0.7 | 0.7 |
| Max | 8.8 | 9.4 | 3.8 | 3.3 | 4.7 | 2.0 |
| Range | 8.3 | 8.8 | 3.6 | 1.4 | 3.9 | 1.3 |
| Median | 3.2 | 4.7 | 1.6 | 2.6 | 2.4 | 1.4 |
| St Dev | 1.9 | 2.3 | 1.1 | 0.3 | 1.2 | 0.3 |
| $\sigma_m$ | 0.2 | 0.2 | 0.1 | 0.0 | 0.1 | 0.0 |
| $C_v$ (%) | 51.3 | 49.5 | 59.7 | 10.6 | 48.4 | 22.7 |
| Correlation Matrix |  |  |  |  |  |  |
| GEO | 1 | -0.19 | -0.22 | -0.30 | 0.35 | 0.37 |
| ARM | -0.19 | 1 | 0.81 | -0.12 | 0.57 | 0.43 |
| AZE | -0.22 | 0.81 | 1 | -0.28 | 0.70 | 0.40 |
| RUS | -0.30 | -0.12 | -0.28 | 1 | -0.44 | 0.04 |
| TUR | 0.35 | 0.57 | 0.70 | -0.44 | 1 | 0.65 |
| WRL | 0.37 | 0.43 | 0.40 | 0.04 | 0.65 | 1 |

**Fig. 2.**
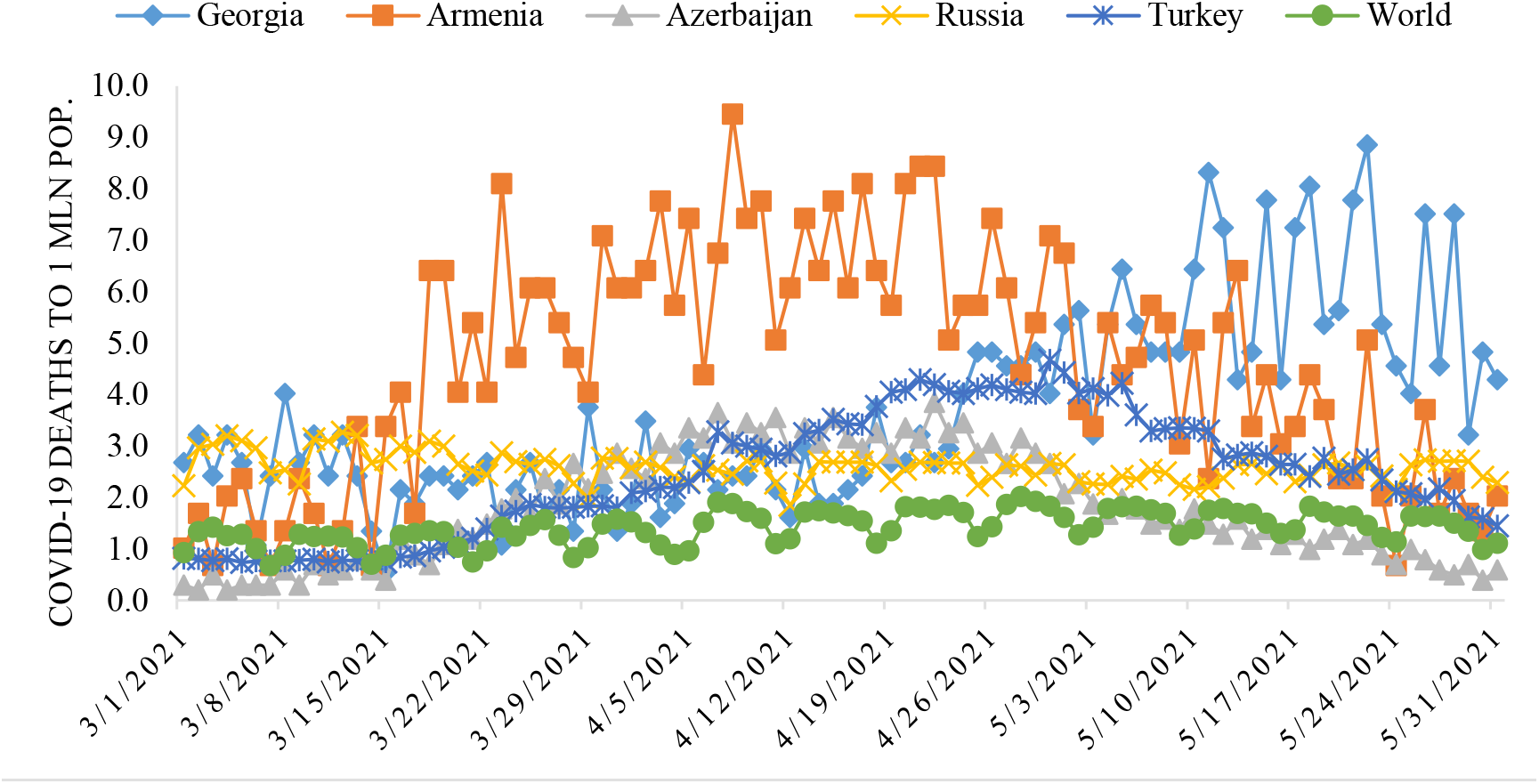
Time-series of deaths cases from Covid-19 to 1 million population in in Georgia, neighboring countries (Armenia, Azerbaijan, Russia, Turkey) and World from March 1, 2021 to May 31, 2021.

- Georgia. Range of change: 0.5-8.8, mean value – 3.7.
- Armenia. Range of change: 0.7-9.4, mean value – 4.6.
- Azerbaijan. Range of change: 0.2-3.8, mean value – 1.8.
- Russia. Range of change: 1.9-3.3, mean value – 2.6.
- Turkey. Range of change: 0.7-4.7, mean value −2.4.
- World. Range of change: 0.7-2.0, mean value – 1.4.

The largest variations in D values were observed in Azerbaijan (C_V_=59.7%), the smallest - in Russia (C_V_=10.6 %).

Significant linear correlation between these countries on D value varies from − 0.44 (pair Russia-Turkey) to +0.81 (pair Armenia-Azerbaijan). Linear correlation between World and these countries besides Russia is significant and varied from +0.37 (pair World-Georgia) to +0.65 (pair World-Turkey). Thus, as in the case of C, except for Russia, in the rest of these countries there are more or less similar trends to world in the variability of mortality from coronavirus (Table 2).

In Table 3 the statistical characteristics of mean monthly values of C and D related to Covid-19 for 156 countries with population ≥ 1 million inhabitants and World in the spring 2021 (normed per 1 million population) is presented.

**Table 3.** The statistical characteristics of mean monthly values of confirmed cases and Deaths cases related to Covid-19 for 156 countries with population ≥ 1 million inhabitant and World from March to May 2021 (normed on 1 mln pop.).

| Month | March |  | April |  | May |  |
| --- | --- | --- | --- | --- | --- | --- |
| Parameter | C | D | C | D | C | D |
| Mean | 124 | 2.0 | 127 | 2.4 | 89 | 1.8 |
| Min | 0 | 0.0 | 0 | 0.0 | 0 | 0.0 |
| Max | 993 | 19.2 | 891 | 23.5 | 1206 | 15.4 |
| Range | 993 | 19.2 | 891 | 23.5 | 1206 | 15.4 |
| St Dev | 182 | 3.4 | 163 | 4.1 | 149 | 2.7 |
| $\sigma_m$ | 15 | 0.27 | 13 | 0.33 | 12 | 0.21 |
| $C_V$ (%) | 147 | 170 | 128 | 168 | 168 | 152 |

As follows from this Table range of change of mean monthly values of C for 156 countries varied from 0 (all months) to 1206 (May). Average value of C for 156 countries varied from 89 (May) to 127 (April). Value of C_V_ changes from 128% (April) to 168% (May).

Range of change of mean monthly values of D for 156 countries varied from 0 (all months) to 23.5 (April). Average value of D for 156 countries varied from 1.8 (May) to 2.4 (April). Value of C_V_ changes from 152% (May) to 170% (March).

Between mean monthly values of Deaths and Confirmed cases related to Covid-19 for 156 countries with population ≥ 1 million inhabitants and World linear correlation and regression are observed (Fig. 3). As follows from Fig. 3 the highest growth rate of the monthly average values of D depending on C was observed in April, the smallest - in May (the corresponding values of the linear regression coefficients).

**Fig. 3.**
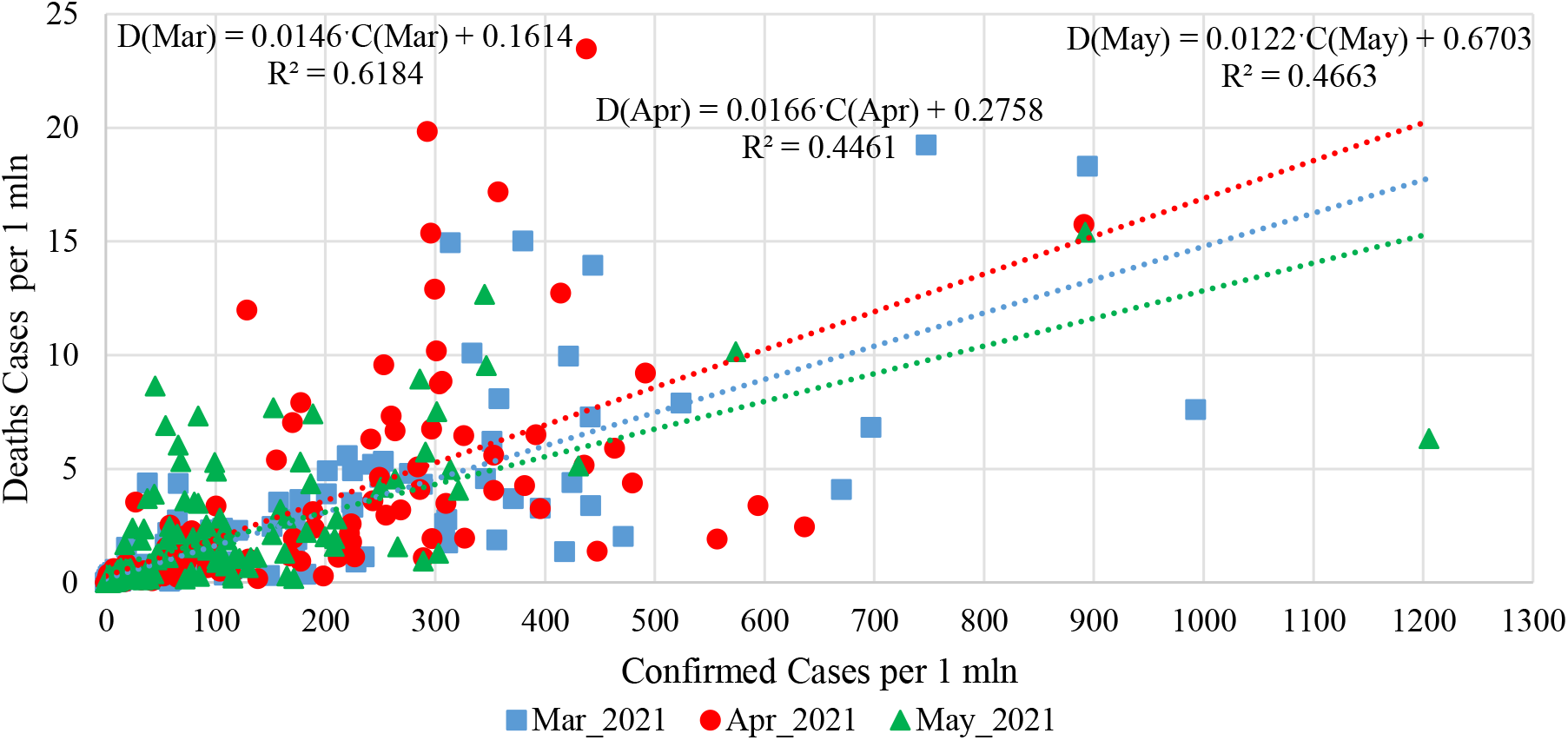
Linear correlation and regression between mean monthly values of Deaths and Confirmed cases related to Covid-19 for 156 countries with population ≥ 1 million inhabitants and world (normed on 1 mln pop.).

In Table 4 data about Covid-19 mean monthly values of infection (C) and deaths (D) cases in spring 2021 (per 1 million population) and ranking of Georgia, Armenia, Azerbaijan, Russia, Turkey and World by these parameters (in brackets) among 156 countries with population ≥ 1 million inhabitant. The corresponding values of the deaths coefficient (DC) are also given here.

**Table 4.**
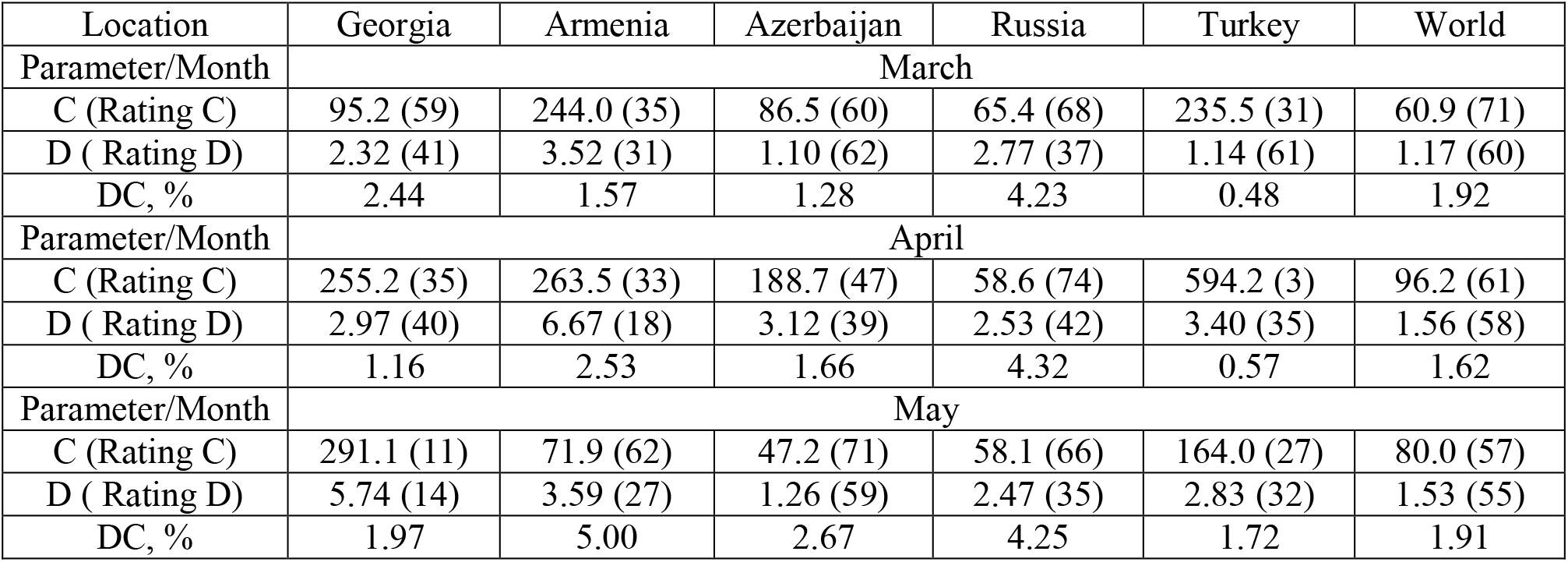
Covid-19 mean monthly values of infection (C) and deaths (D) cases in spring 2021 (per 1 million population) and ranking of Georgia, Armenia, Azerbaijan, Russia, Turkey and World by these parameters (in brackets) among 156 countries with population ≥ 1 million inhabitant, and correspondent values of deaths coefficient (DC).

In particular, as follows from this Table, mean monthly values of C for 5 country changes from 47.2 (Azerbaijan, May, 71 place between 156 country and world in this month) to 594.2 (Turkey, April, 3 place between 156 country and world in this month).

Mean monthly values of D for 5 country changes from 1.10 (Azerbaijan, March, 62 place between 156 country and World in this month) to 6.67 (Armenia, April, 18 place between 156 country and World in this month).

Among 156 countries with population ≥ 1 million inhabitants and World in May 2021 Georgia was in the 11 place on new infection cases (291.1) and in the 14 place on Death (5.74).

Mean monthly values of DC for 5 country changes from 0.48% (Turkey, March) to 5.00 (Armenia, May).

Note that the mean values of DC (%) in spring 2021 are: Georgia – 1.73, Armenia – 2.46, Azerbaijan – 1.70, Russia – 4.26, Turkey – 0.74, World – 1.80 (according to data from Table 1 and 2).

Fig. 4. The rate of new infection of the population of Georgia, Armenia, Azerbaijan, Russia and, Turkey with Covid-19 according to traffic light system from March 1, 2021 to May 31, 2021.

So, in the most unfavorable red zone, Georgia was from March 31 to May 31, Armenia – from March 8 to May 13, Azerbaijan - from March 28 to May 7, Turkey – from March 4 to May 29. Russia, as a whole, was also in the red zone (average sum 14 day values of C per 100000 populations = 87, and I was more than 4%).

**Fig. 4.**
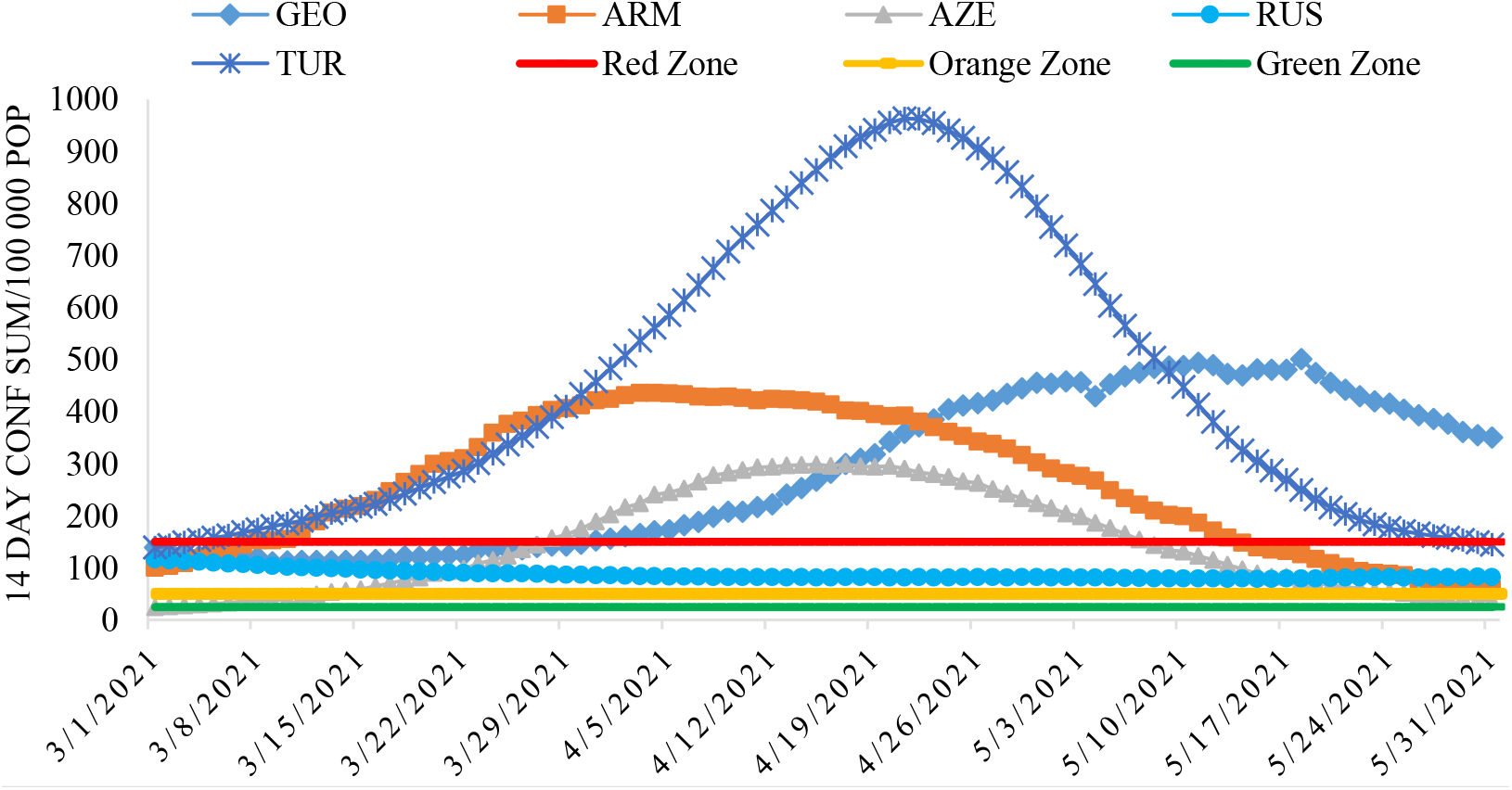
demonstrated the rate of new infection of the population of Georgia, Armenia, Azerbaijan, Russia and Turkey with Covid-19 according to traffic light system in the spring 2021.

### 3.2 Comparison of daily death from Covid-19 in Georgia with daily mean death in 2015-2019 from March 1, 2021 to May 31, 2021

In Fig. 5 data of the daily death from Covid-19 in Georgia from October 1, 2020 to May 31, 2021 in comparison with daily mean death in 2015-2019. The daily mean death in 2015-2019 in different months are: October – 123, November – 135, December – 144, January – 155, February – 146, March – 141, April – 137, May – 131.

**Fig. 5.**
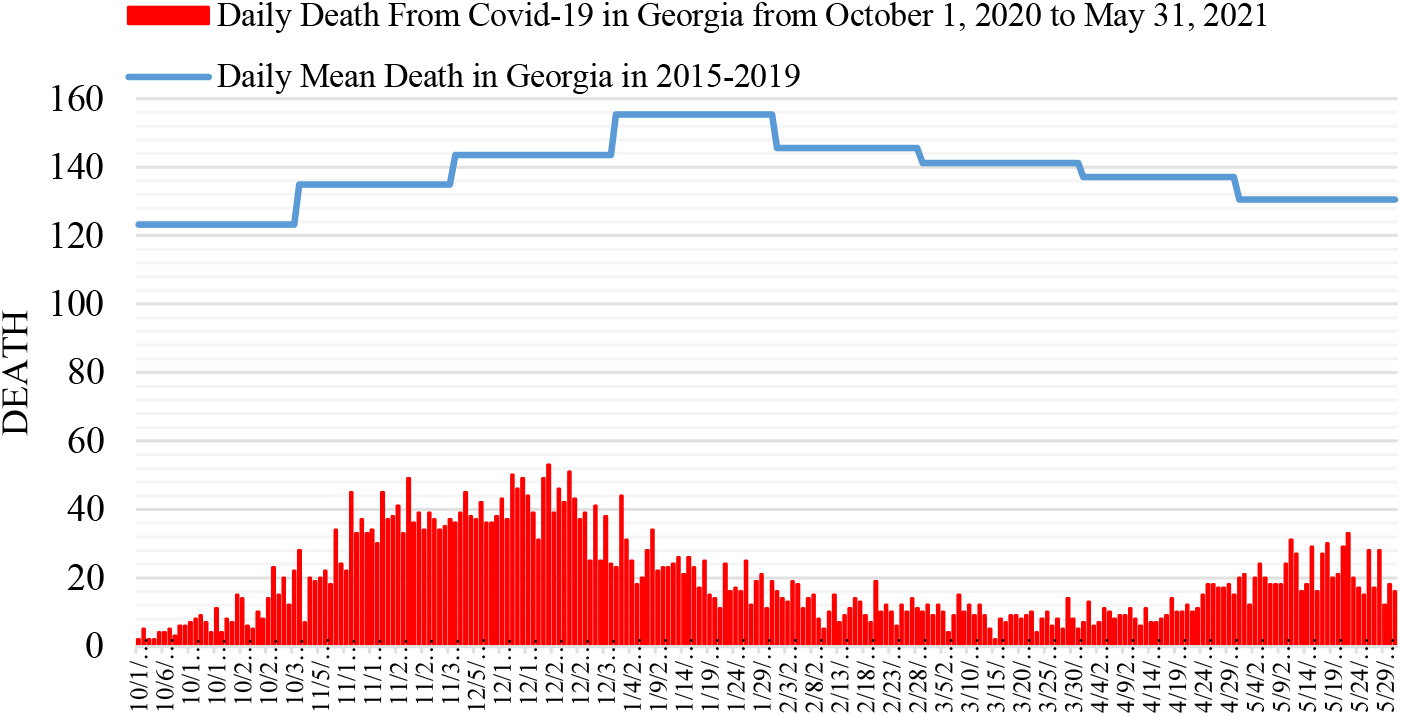
Daily death from Covid-19 in Georgia from October 1, 2020 to May 31, 2021 in comparison with daily mean death in 2015-2019.

The daily death from Covid-19 from March 1, 2021 to May 31, 2021 changes from 2 to 33.

Fig. 6 shows, that the largest share value of D in spring 2021 from mean death in 2015-2019 was 25.3 % (22.05.2021), the smallest 1.42 % (15.03.2021).

**Fig. 6.**
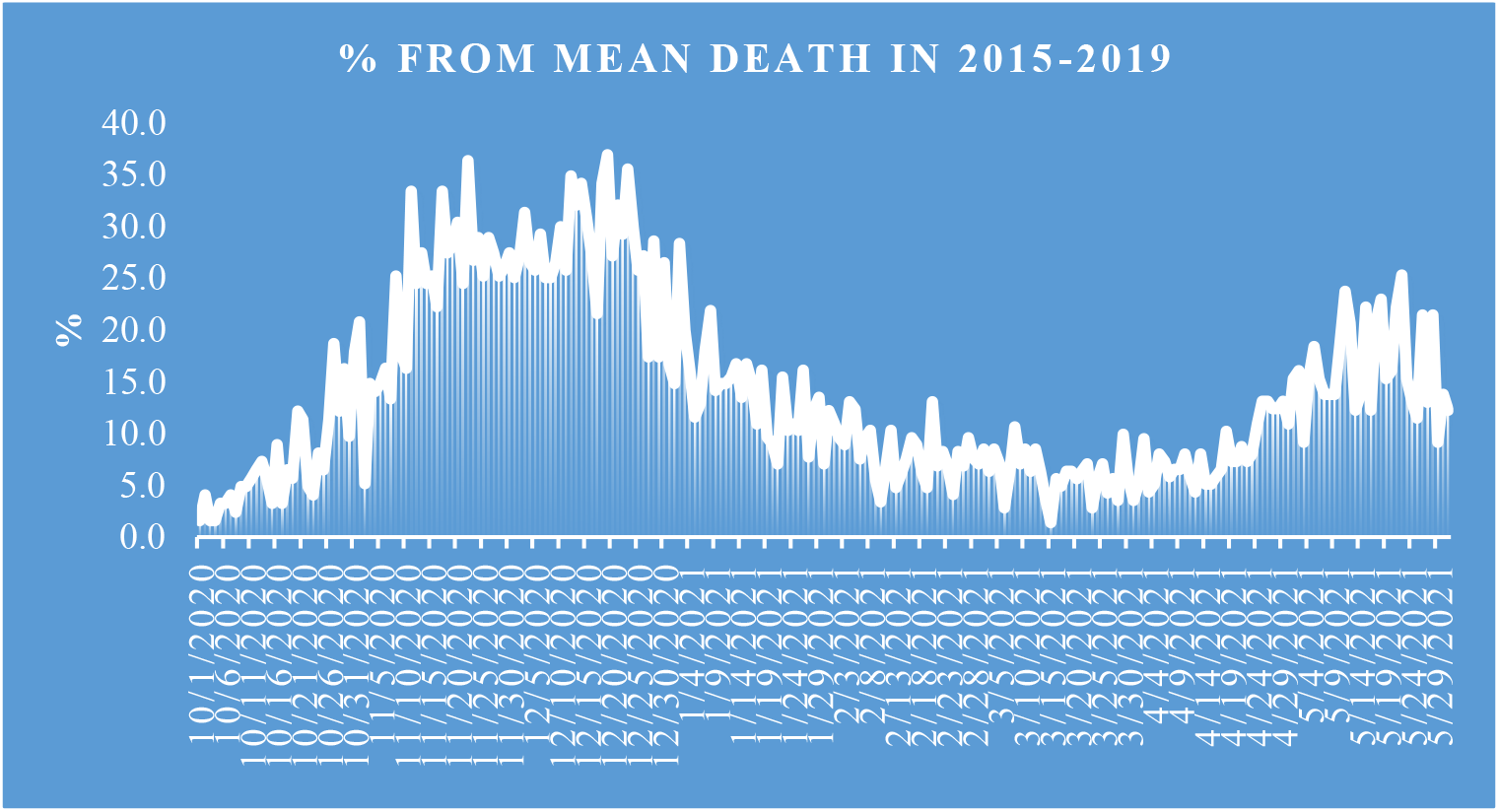
Share value of D from mean death in 2015-2019.

### 3.3 The statistical analysis of the daily and decade data associated with New Coronavirus COVID-19 pandemic in spring 2021

Results of the statistical analysis of the daily and decade data associated with New Coronavirus COVID-19 pandemic in Georgia from March 1, 2021 to May 31, 2021 in Tables 5-7 and Fig. 7 – 18 are presented.

**Table 5.** Statistical characteristics of the daily data associated with coronavirus COVID-19 pandemic of confirmed, recovered, deaths cases and infection rate of the population of Georgia from 01.03.2021 to 31.05.2021.

| Parameter | Confirmed | Recovered | Deaths | Infection Rate, % |
| --- | --- | --- | --- | --- |
| Mean | 796 | 680 | 14 | 3.3 |
| Min | 133 | 91 | 2 | 0.9 |
| Max | 2171 | 2038 | 33 | 8.0 |
| Range | 2038 | 1947 | 31 | 7.2 |
| St Dev | 482 | 464 | 7 | 1.7 |
| $\sigma_m$ | 51 | 49 | 0.7 | 0.2 |
| $C_v$ (%) | 60.6 | 68.2 | 51.3 | 52.7 |

**Fig. 7.**
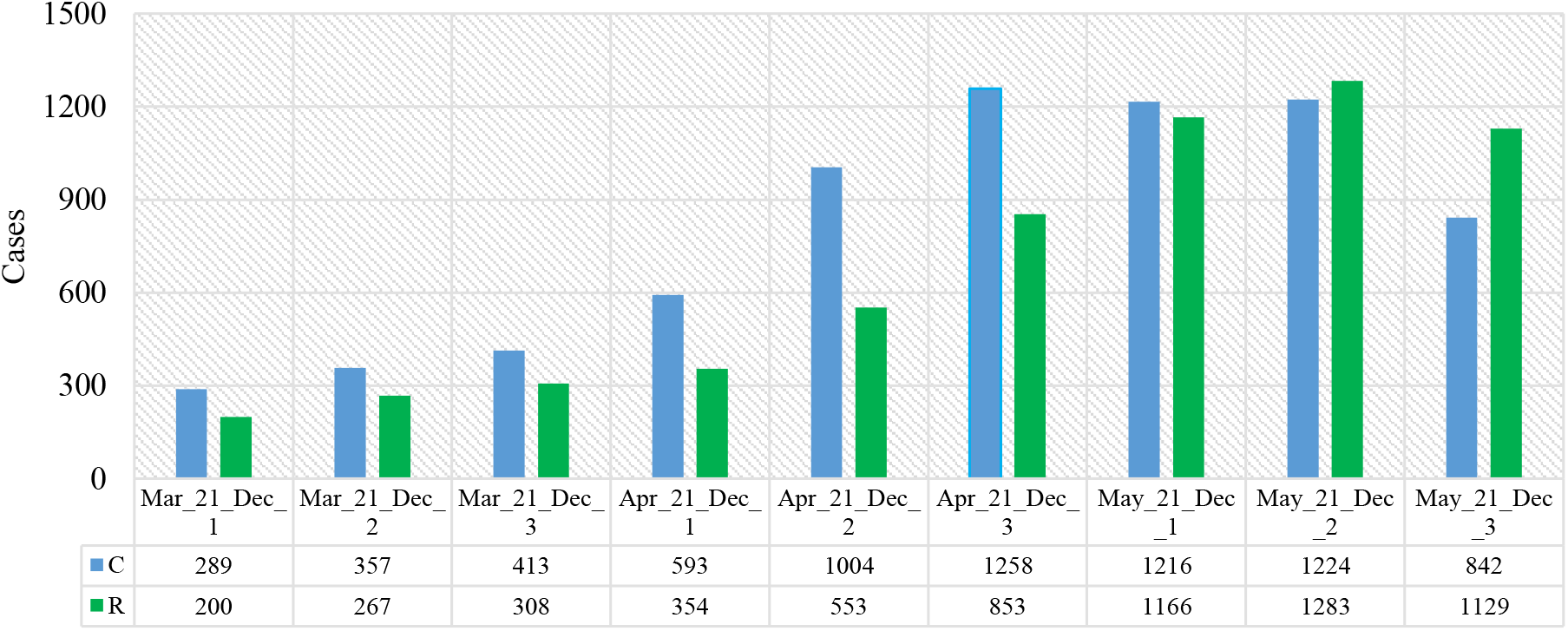
Mean values of confirmed and recovered coronavirus-related cases in different decades of months in Georgia from March 2021 to May 2021.

The mean and extreme values of the studied parameters are as follows (Table 5): C - mean - 796, range: 133 - 2171; R - mean - 680, range: 91 - 2038; D - mean - 14, range: 2 - 33; I (%) - mean – 3.3, range: 0.9 – 8.0. All studied parameters are subject to notable variations: 51.3 % (D) ≤C_**V**_ ≤68.2%( R).

Mean decade values of confirmed and recovered coronavirus-related cases varies within the following limits (Fig. 7): C – from 289 (1 Decade of March 2021) to 1258 (3 Decade of April 2021); R – from 200 (1 Decade of March 2021) to 1283 (2 Decade of May 2021).

Mean decade values of deaths coronavirus-related cases (Fig. 8) varies from 8 (2 and 3 Decades of March) to 24 (2 Decade of May).

**Fig. 8.**
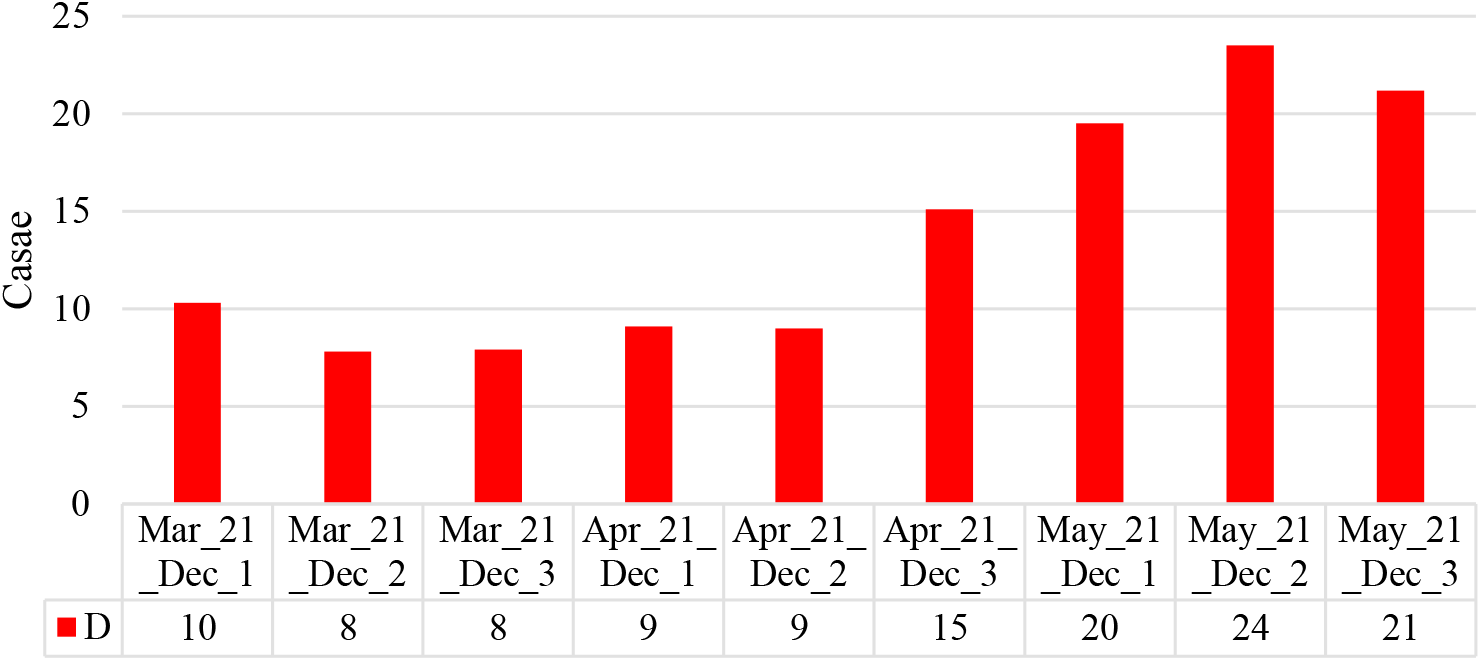
Mean values of deaths coronavirus-related cases in different decades of months in Georgia from March 2021 to May 2021.

Mean decade values of infection rate coronavirus-related cases (Fig. 9) varies from 1.70 % (1 Decade of March) to 6.54 % (1 Decade of May).

**Fig. 9.**
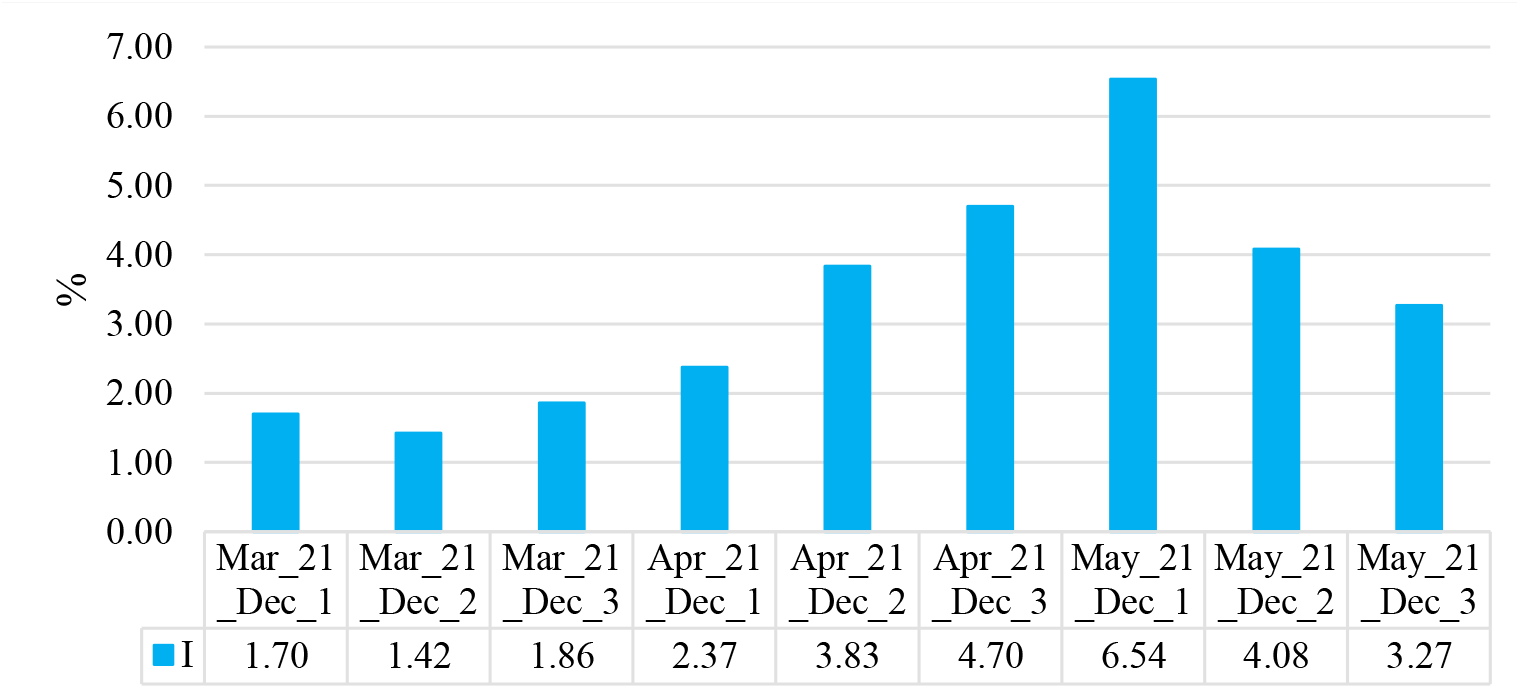
Mean values of infection rate coronavirus-related cases in different decades of months in Georgia from March 2021 to May 2021.

Time changeability of the daily values of C, R, D and I are satisfactorily described by the tenth order polynomial (Table 6, Fig. 10-12). For clarity, the data in Fig. 11 are presented in relative units (%) in relation to their average values

**Table 6.** Form of the equations of the regression of the time changeability of the daily values of C, R and D from March 01, 2021 to May 31, 2021 in Georgia. The level of significance of R^2^ is not worse than 0.001.

| Parameter | Confirmed | Recovered | Deaths | Infection Rate |
| --- | --- | --- | --- | --- |
| Regression | Tenth Order Polynomial |  |  |  |
| $R^2$ | 0.629 | 0.790 | 0.739 | 0.821 |
| $K_{DW}$ | 1.99 | 2.17 | 2.23 | 0.917 |
| Autocorrelation of residual components | Absent | Absent | Absent | Positive |

**Fig. 10.**
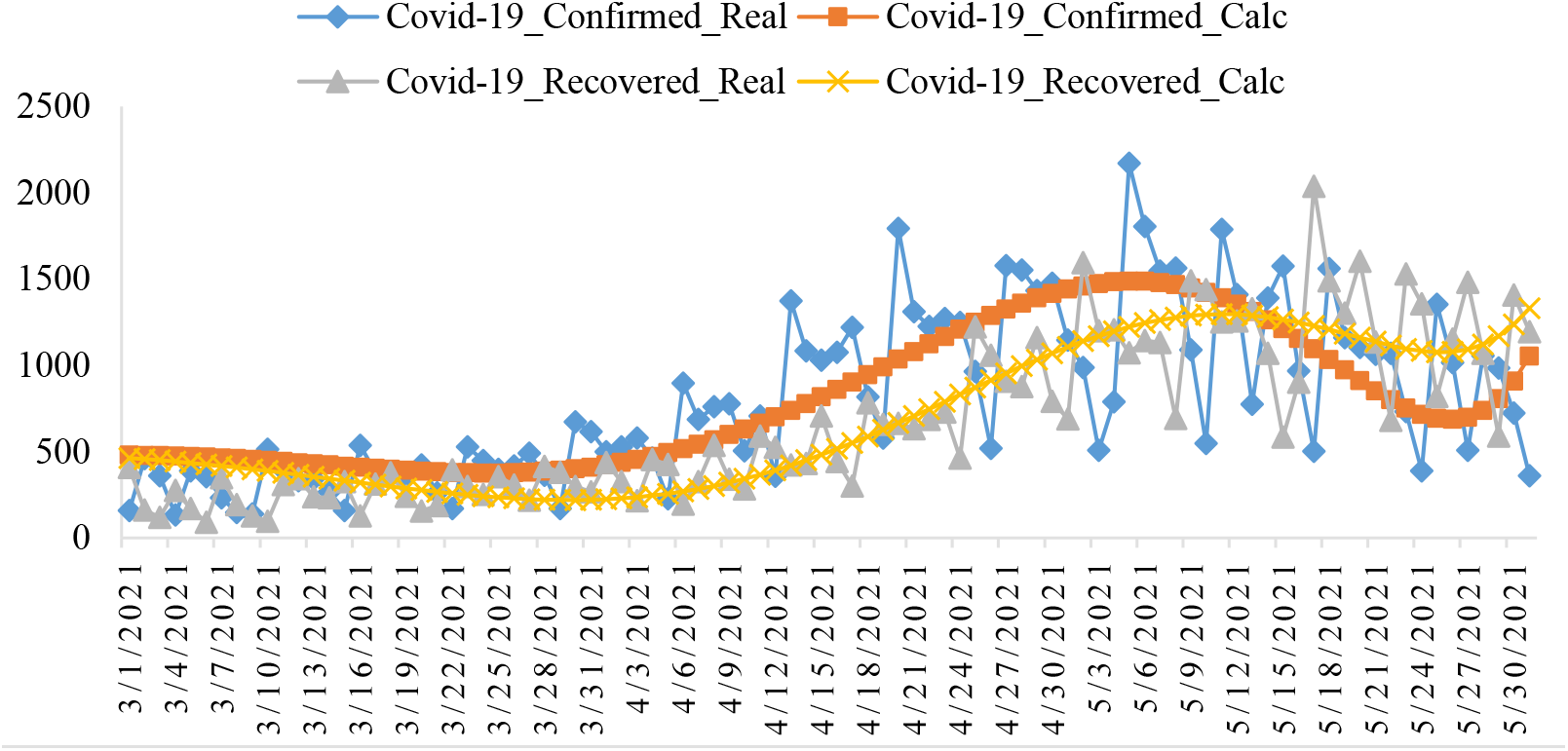
Changeability of the real and calculated daily values of coronavirus confirmed and recovered cases from March 01, 2021 to May 31, 2021 in Georgia.

**Fig. 11.**
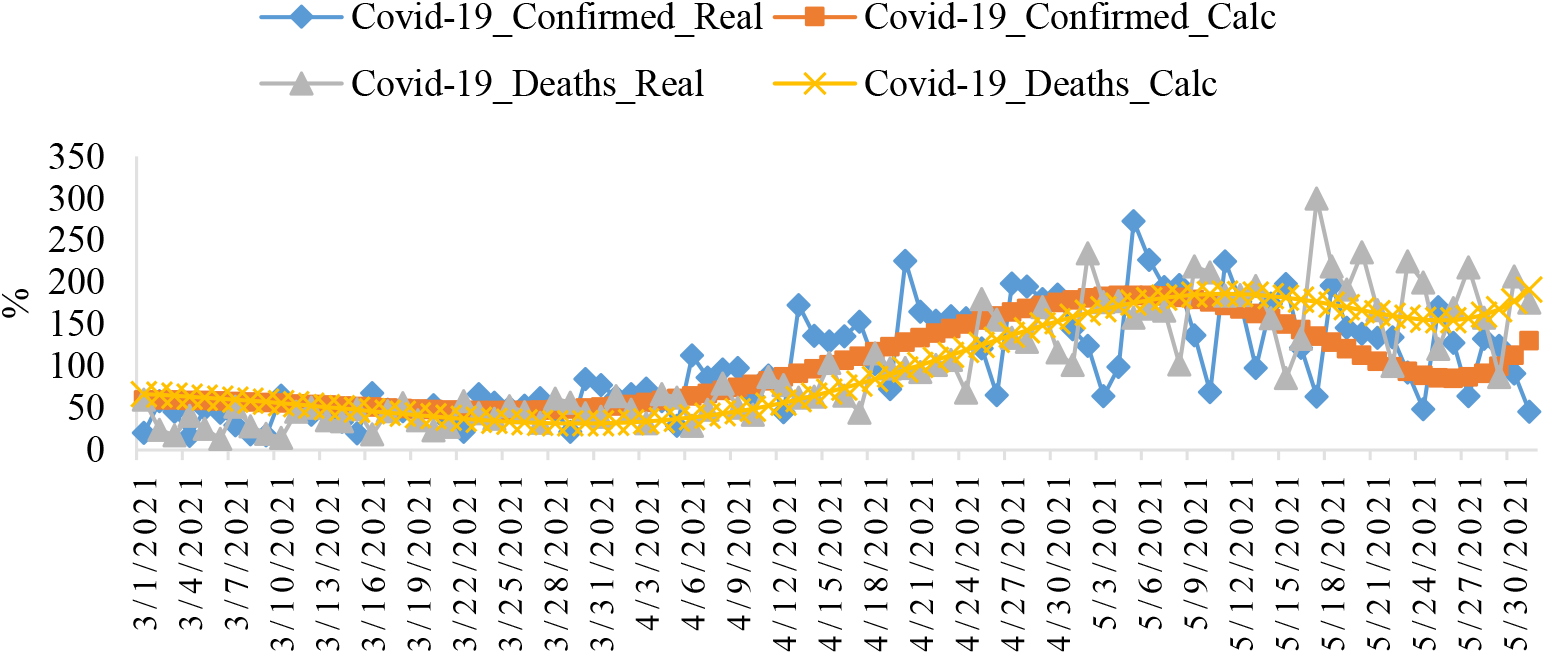
Changeability of the real and calculated daily values of coronavirus-related confirmed and deaths cases from March 01, 2021 to May 31, 2021 in Georgia.

**Fig. 12.**
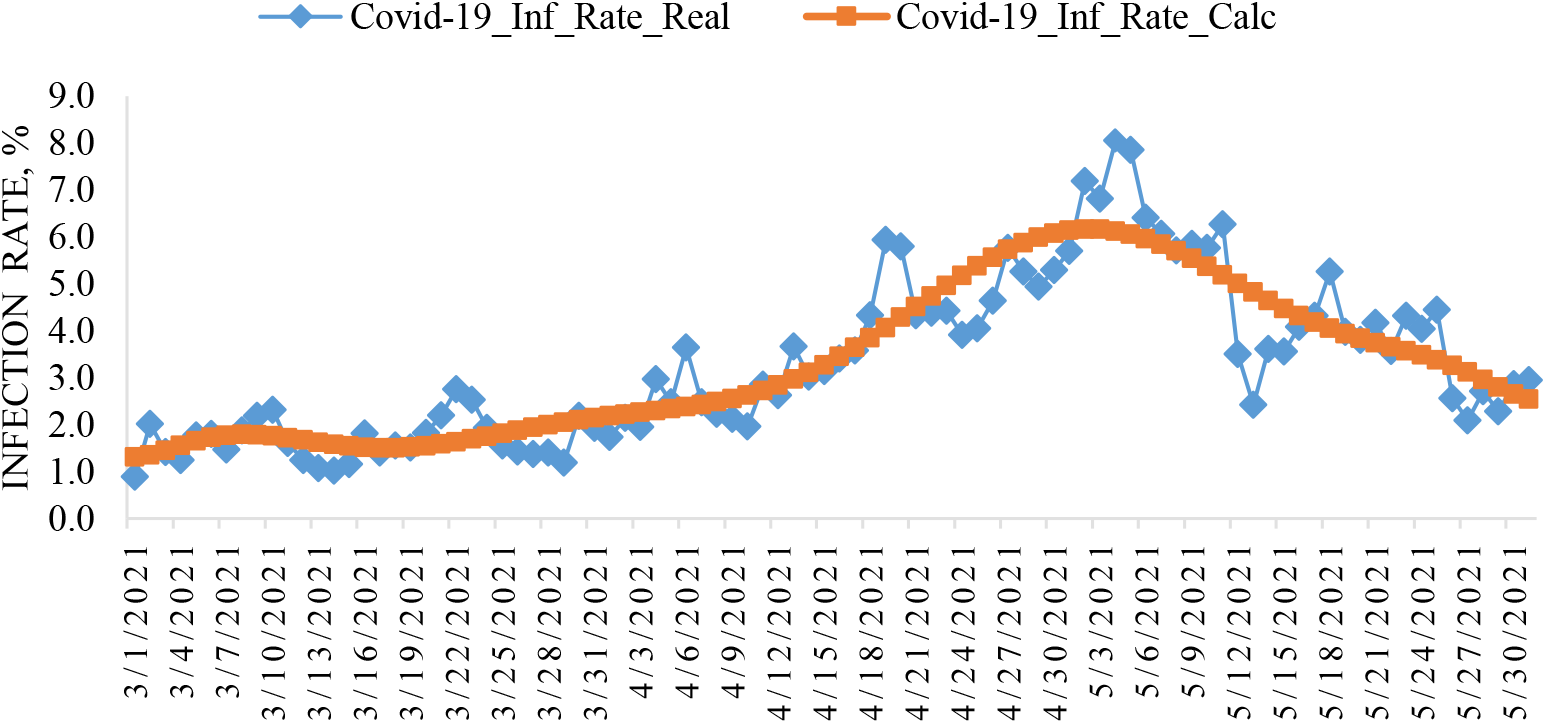
Changeability of the real and calculated daily values of coronavirus Infection Rate from March 01, 2021 to May 31, 2021 in Georgia.

Note that from Fig. 10 and 11, as in [8], clearly show the shift of the time series values of R and D in relation to C.

In Fig. 13-15 data about mean values of speed of change of confirmed, recovered, deaths coronavirus-related cases and infection rate in different decades of months in spring 2021 are presented.

**Fig. 13.**
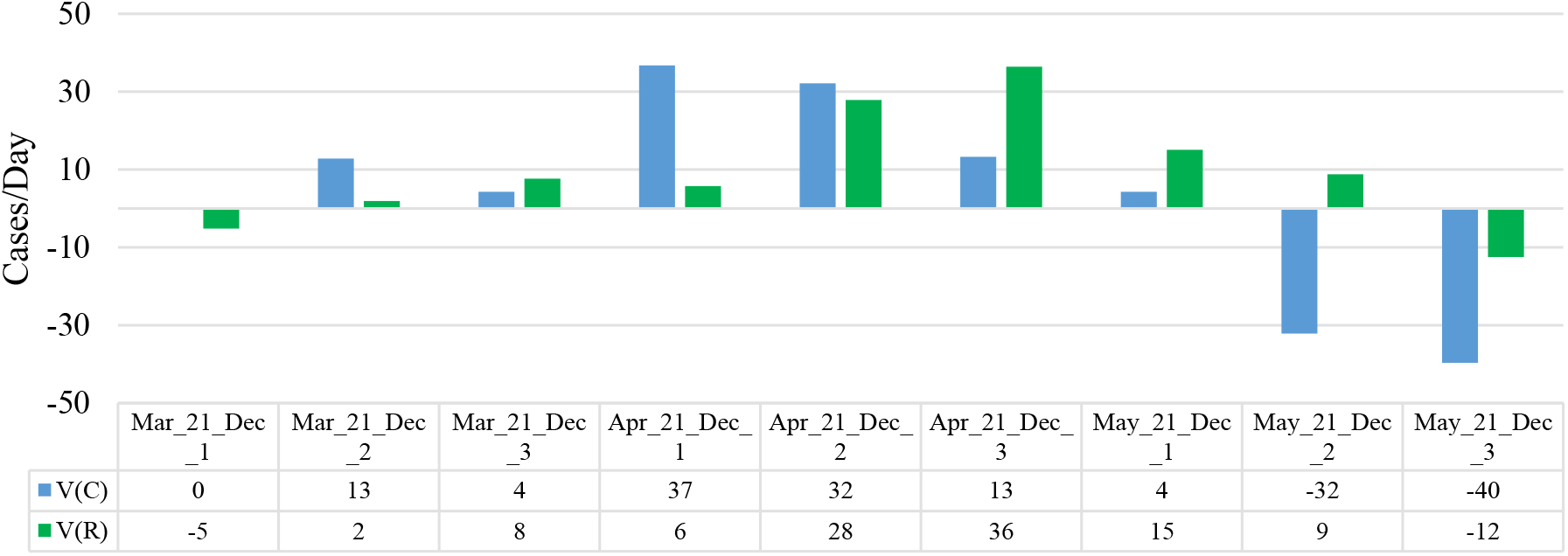
Mean values of speed of change of confirmed and recovered coronavirus-related cases in different decades of months in Georgia from March 2021 to May 2021.

**Fig. 14.**
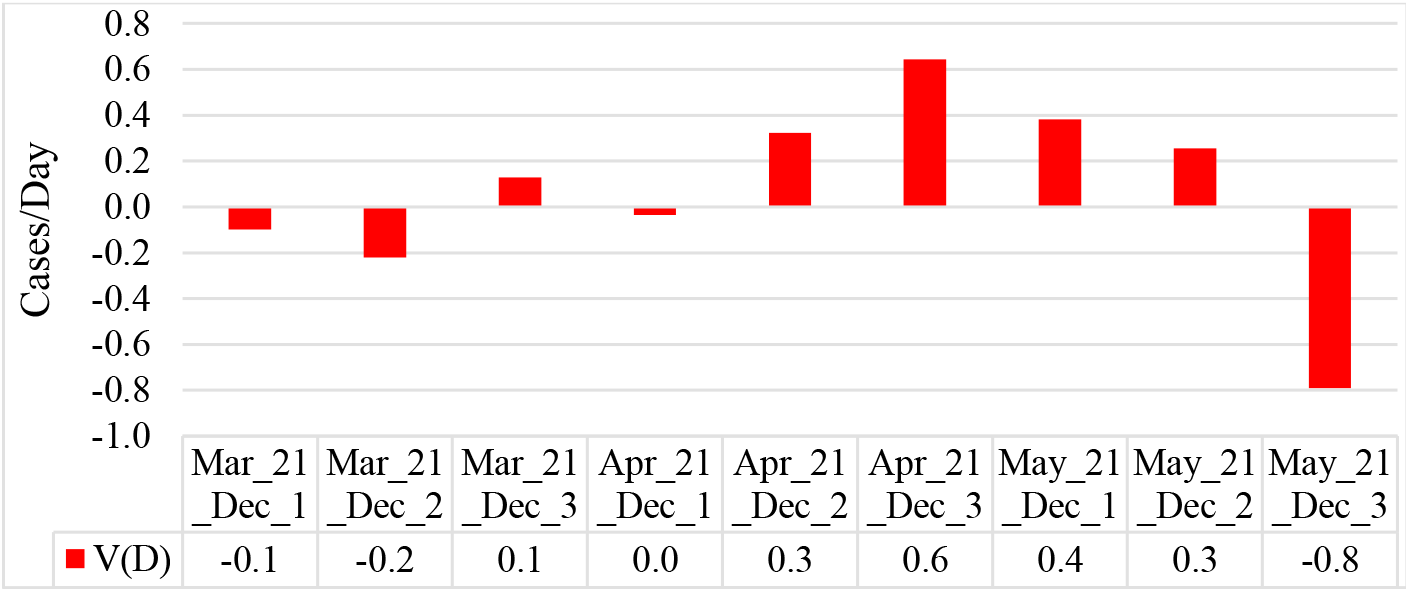
Mean values of speed of change of deaths coronavirus-related cases in different decades of months in Georgia from March 2021 to May 2021.

**Fig. 15.**
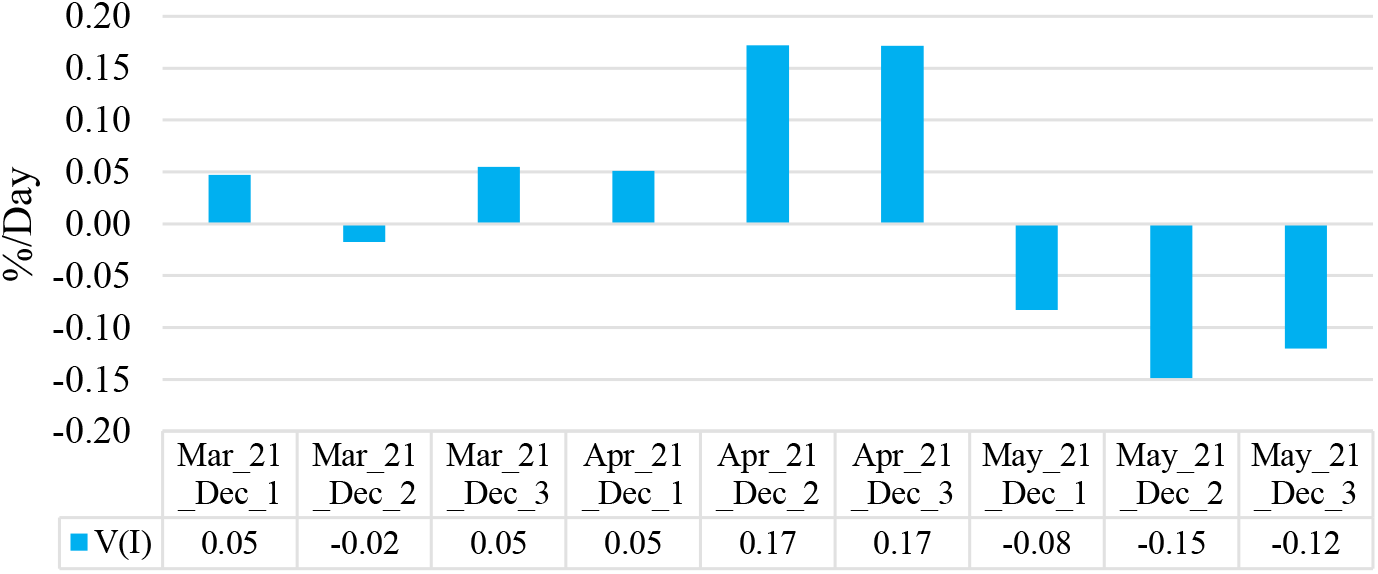
Mean values of speed of change of coronavirus infection rate in different decades of months in Georgia from March 2021 to May 2021.

Maximum mean decade values of investigation parameters are following: V(C) = +37 cases/day (1 Decade of April), V(R) = +36 cases/day (3 Decade of April), V(D) = +0.6 cases/day (3 Decade of April), V(I) = + 0.17 %/ day (2 and 3 decades of April). Min mean decade values of investigation parameters are following: V(C) = −40 cases/day (3 Decade of May), V(R) = −12 cases/day (3 Decade May), V(D) = −0.8 cases/day (3 Decade May), V(I) = −0.15 %/day (2 Decade of May).

Data about mean monthly values of C, R, D, I and its speed of change in Georgia in spring 2021 in Table 7 are presented. As follows from this Table there was an increase in average monthly values of C, R, D and I from March to May. As for the values of V(C), V(R), V(D) and V(I), their growth was observed in April in relation to March. In May, there was a decrease in all these parameters in relation to the two previous months. That is, some tendencies have emerged to improve the epidemiological situation in Georgia.

**Table 7.** Mean monthly values of C, R, D, I and its speed of change in Georgia from March 2021 to May 2021.

| Parameter | C | R | D | I, % | $V(C)$ | $V(R)$ | $V(D)$ | $V(I)$ |
| --- | --- | --- | --- | --- | --- | --- | --- | --- |
| Mar_2021 | 355 | 260 | 9 | 1.67 | 5.7 | 1.9 | -0.1 | 0.03 |
| Apr_2021 | 952 | 587 | 11 | 3.63 | 27.3 | 23.3 | 0.3 | 0.13 |
| May_2021 | 1086 | 1190 | 21 | 4.59 | -22.5 | 3.8 | -0.1 | -0.12 |

In Fig. 16 data about connection of 14-day moving average of deaths cases due to COVID-19 in Georgia with 14-day moving average of infection rate from December 18, 2020 until May 31, 2021 are presented. As follows from Fig. 16, in general, with an increase of the infection rate is observed increase of deaths cases. However, in May 2021, the average death rate from coronavirus was almost the same as in January 2021 (21.4 and 21.7, respectively). Although the average values of I in May were significantly lower than in January (4.3% and 7.7%, respectively, Fig. 17). This fact needs special analysis.

**Fig. 16.**
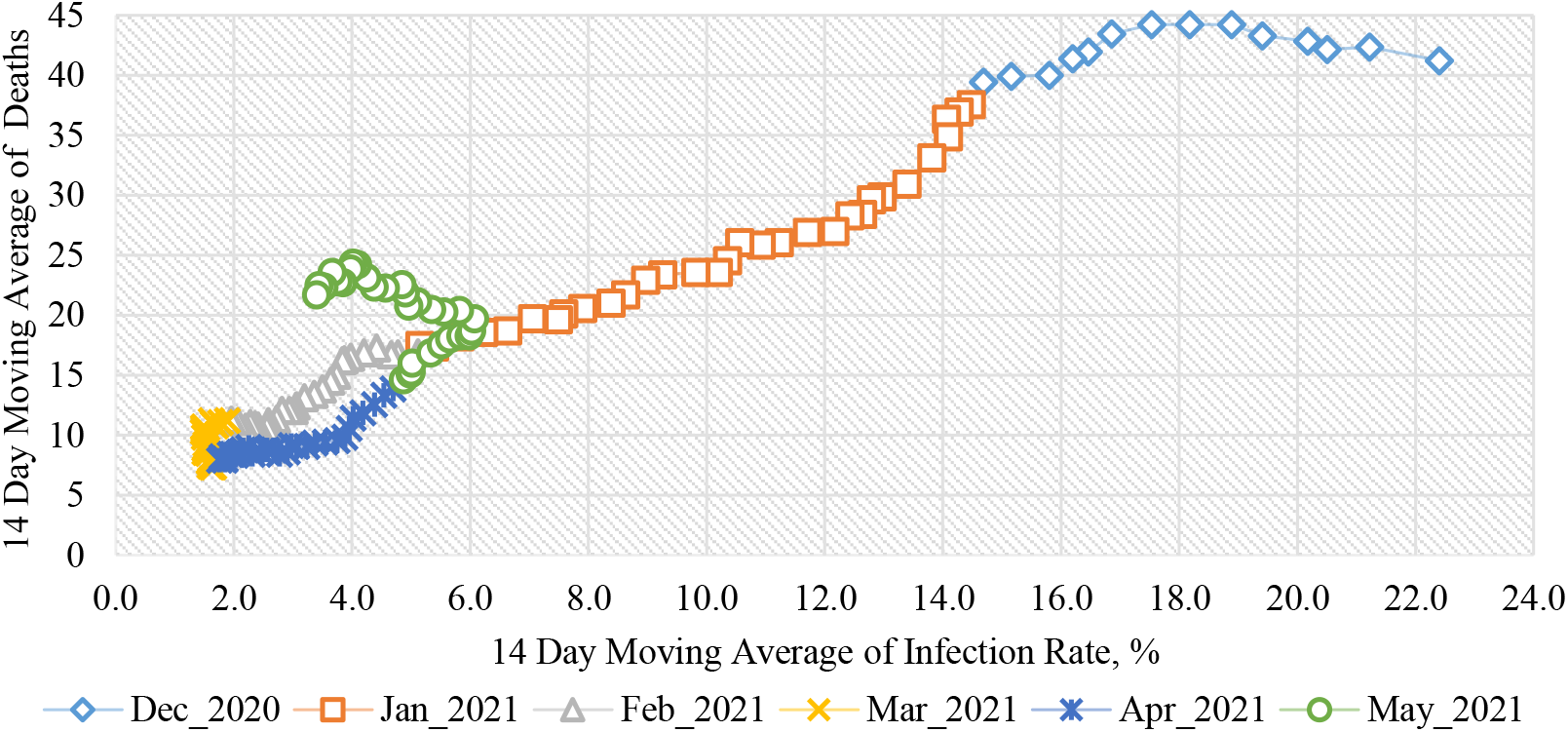
Connection of 14-day moving average of deaths cases due to COVID-19 in Georgia with 14-day moving average of infection rate from December 18, 2020 until May 31, 2021.

**Fig. 17.**
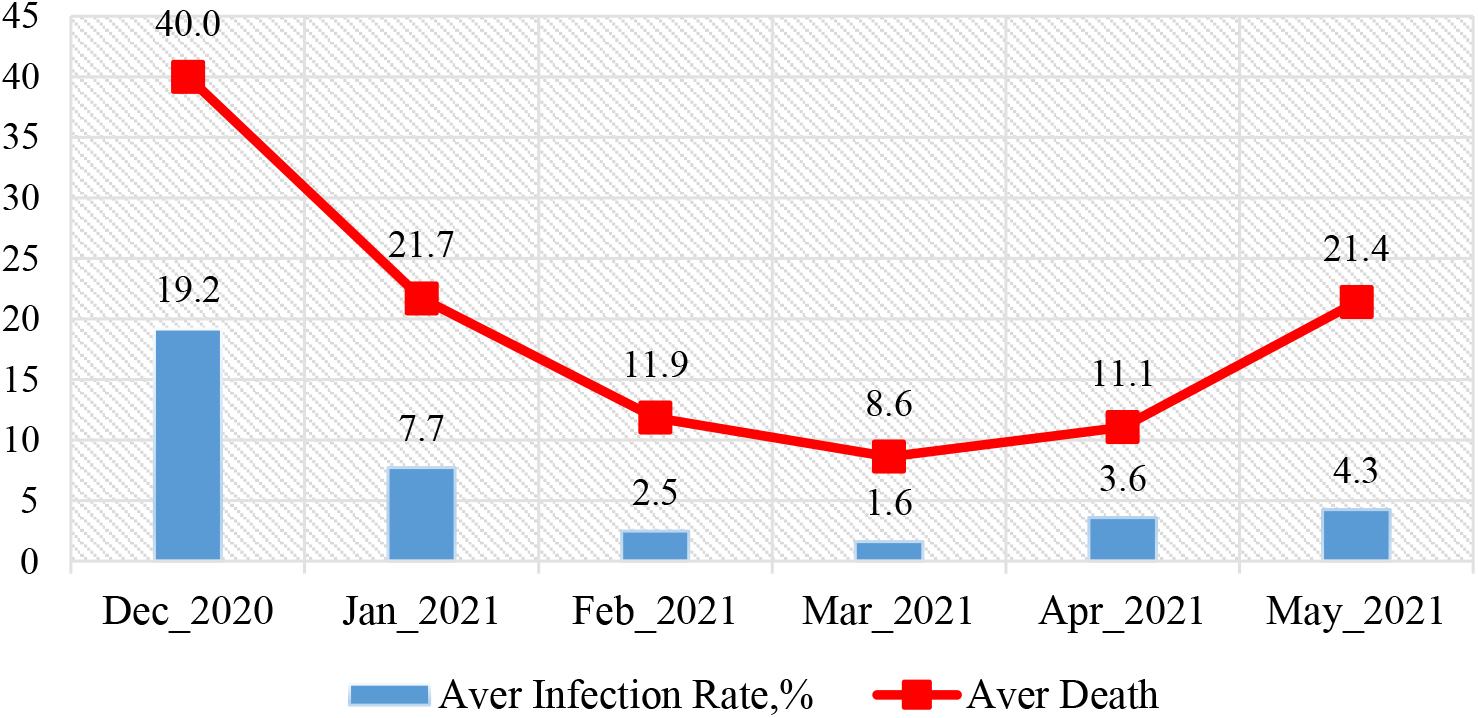
Mean monthly values of COVID-19 infection rate and deaths cases in Georgia from December 2020 to May 2021.

Using the data in Fig. 17, a linear regression graph between the monthly mean values of D and I is obtained (Fig. 18). As follows from Fig. 18, a high level of linear correlation between these parameters is observed. This Fig. also clearly demonstrates the anomaly high mortality from coronavirus in May 2021 with relatively low value of infection rate.

**Fig. 18.**
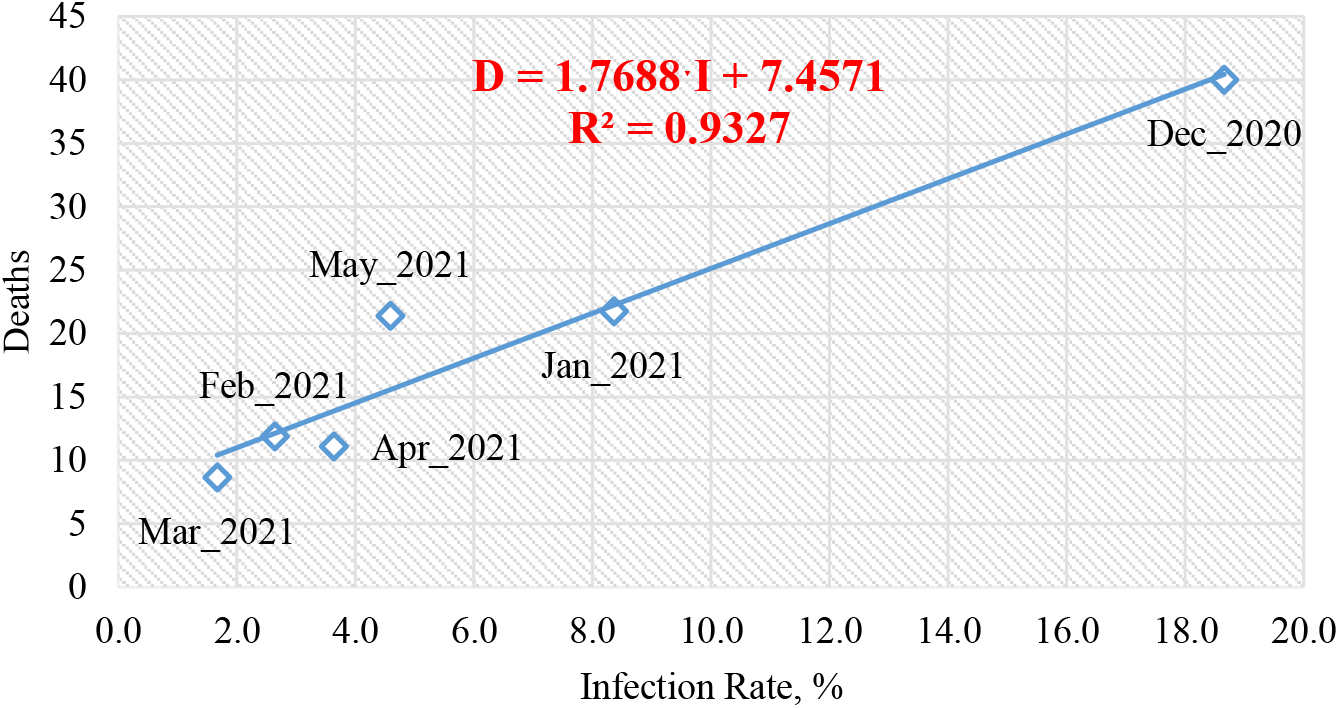
Linear correlation and regression between the mean monthly values of COVID-19 infection rate and deaths cases in Georgia from December 2020 to May 2021.

As noted in [8] and above (Fig. 10 and 11), there is some time-lag in the values of the time series R and D with respect to C. An estimate of the values of this time-lag is given below (Fig. 19).

**Fig. 19.**
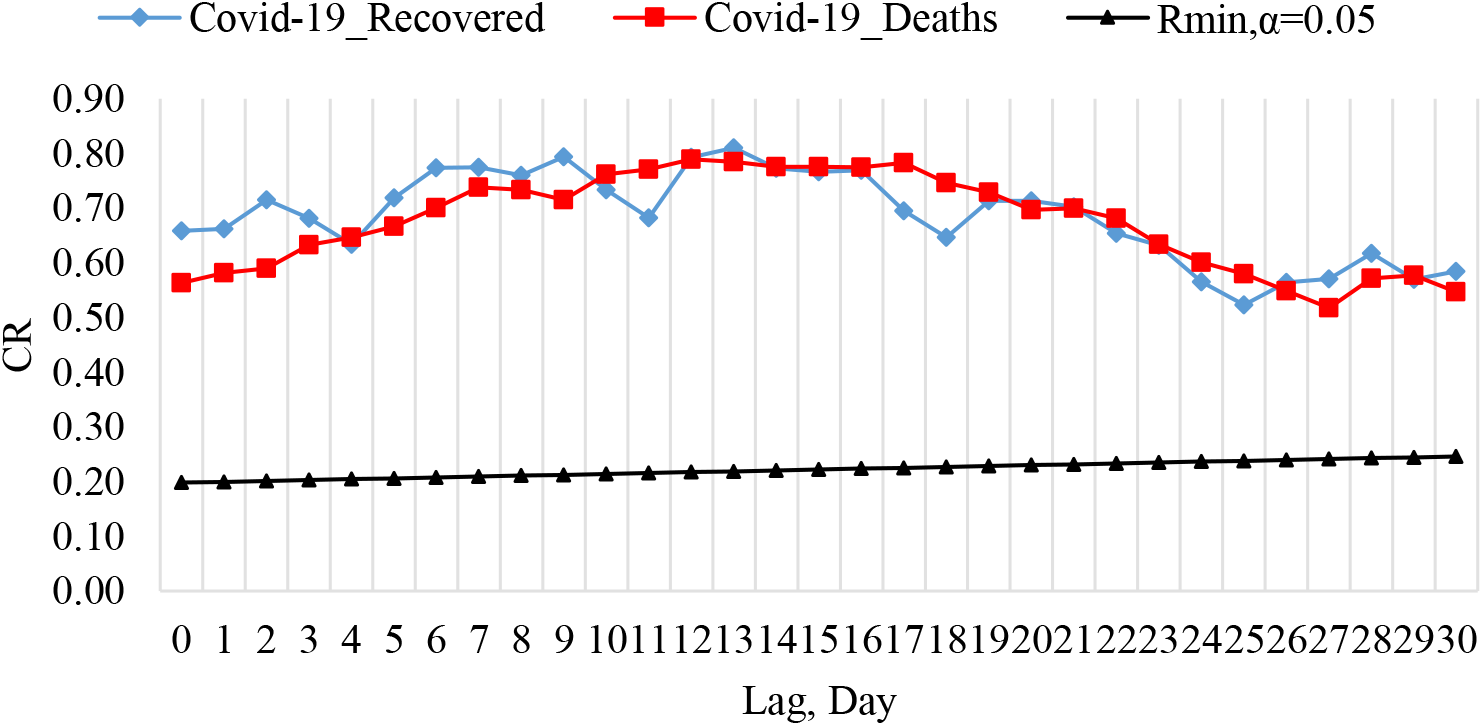
Coefficient of cross-correlations between confirmed COVID-19 cases (normed to tests number) with recovered and deaths cases in Georgia from 01.03.2021 to 31.05.2021.

Cross-correlations analysis between confirmed COVID-19 cases with recovered and deaths cases shows, that the maximum effect of recovery is observed 9 and 13 days after infection, and deaths - after 12-17 days (Fig. 19). Earlier it was found that the maximum effect of recovery is observed 13-14 days after infection, and deaths - after 13-14 and 17-18 days [8]. In general, both recoveries and deaths can occur soon enough or long after infection. These effects will be refined as new data accumulates.

### 3.4 Comparison of real and calculated prognostic daily and two weekly data related to the New Coronavirus COVID-19 pandemic in Georgia in spring 2021

In Fig. 20-23 and Table 8 examples of comparison of real and calculated prognostic daily and mean two weekly data related to the COVID-19 coronavirus pandemic in Georgia in spring 2021are presented. Note that the results of the analysis of two-week forecasting of the values of C, D and I, information about which was regularly sent to the National Center for Disease Control & Public Health of Georgia and posted on the Facebook page https://www.facebook.com/Avtandil1948/.

**Fig. 20.**
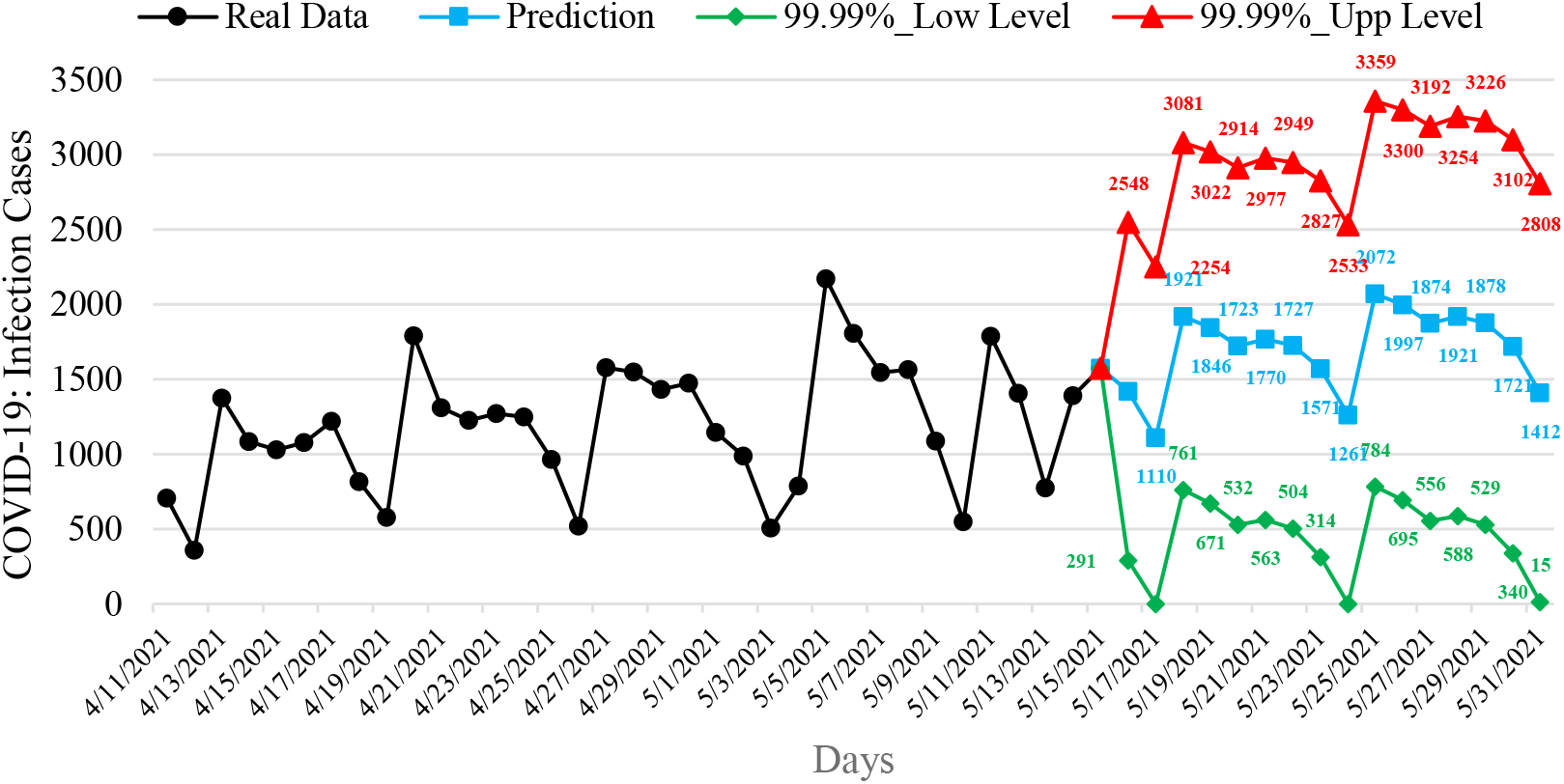
Example of Interval Prediction of COVID-19 Infection Cases in Georgia from 16.05.2021 to 31.05.2021.

**Fig. 21.**
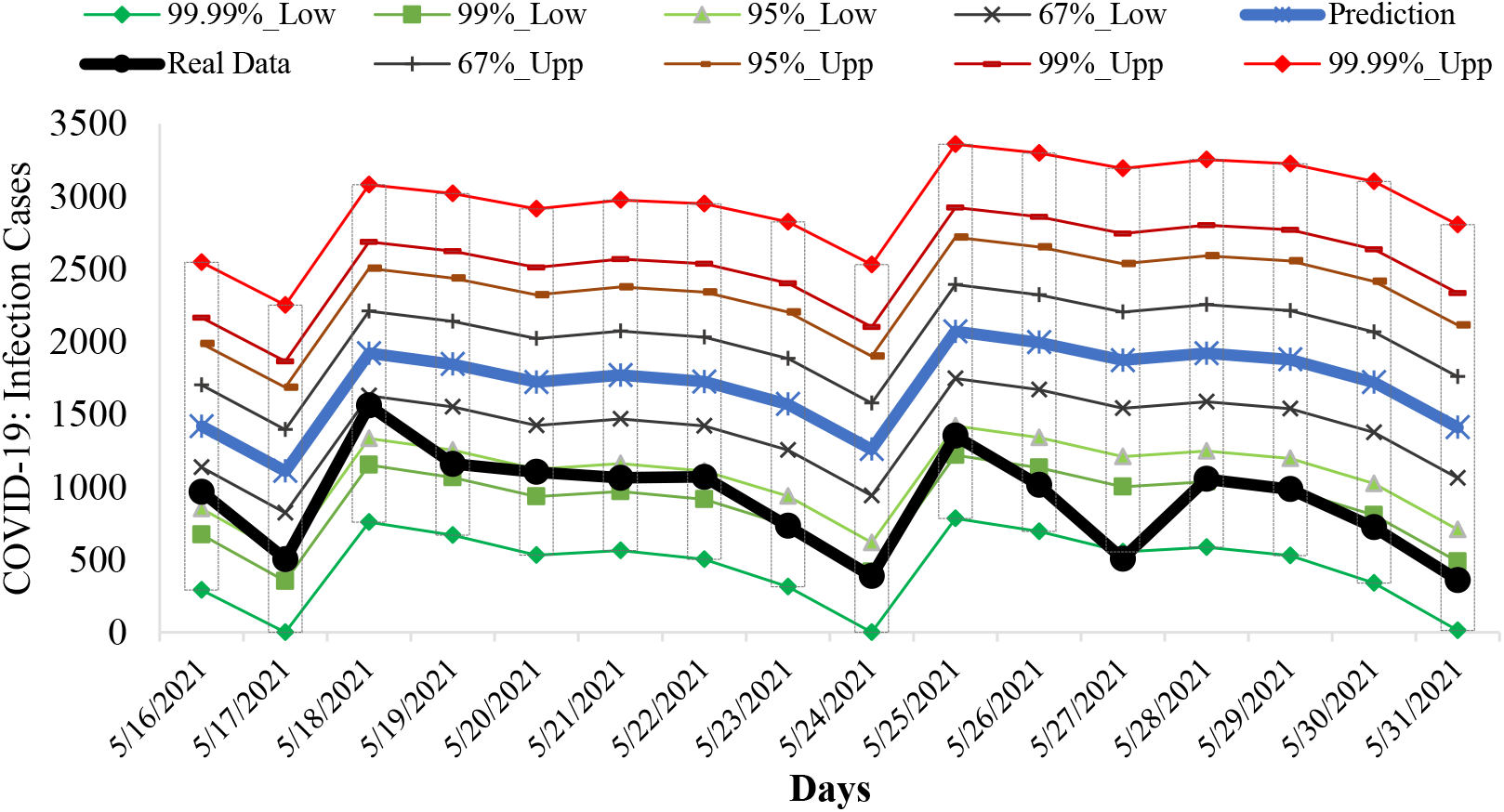
Example of Verification of Interval Prediction of COVID-19 Infection Cases in Georgia from 16.05.2021 to 31.05.2021.

**Fig. 22.**
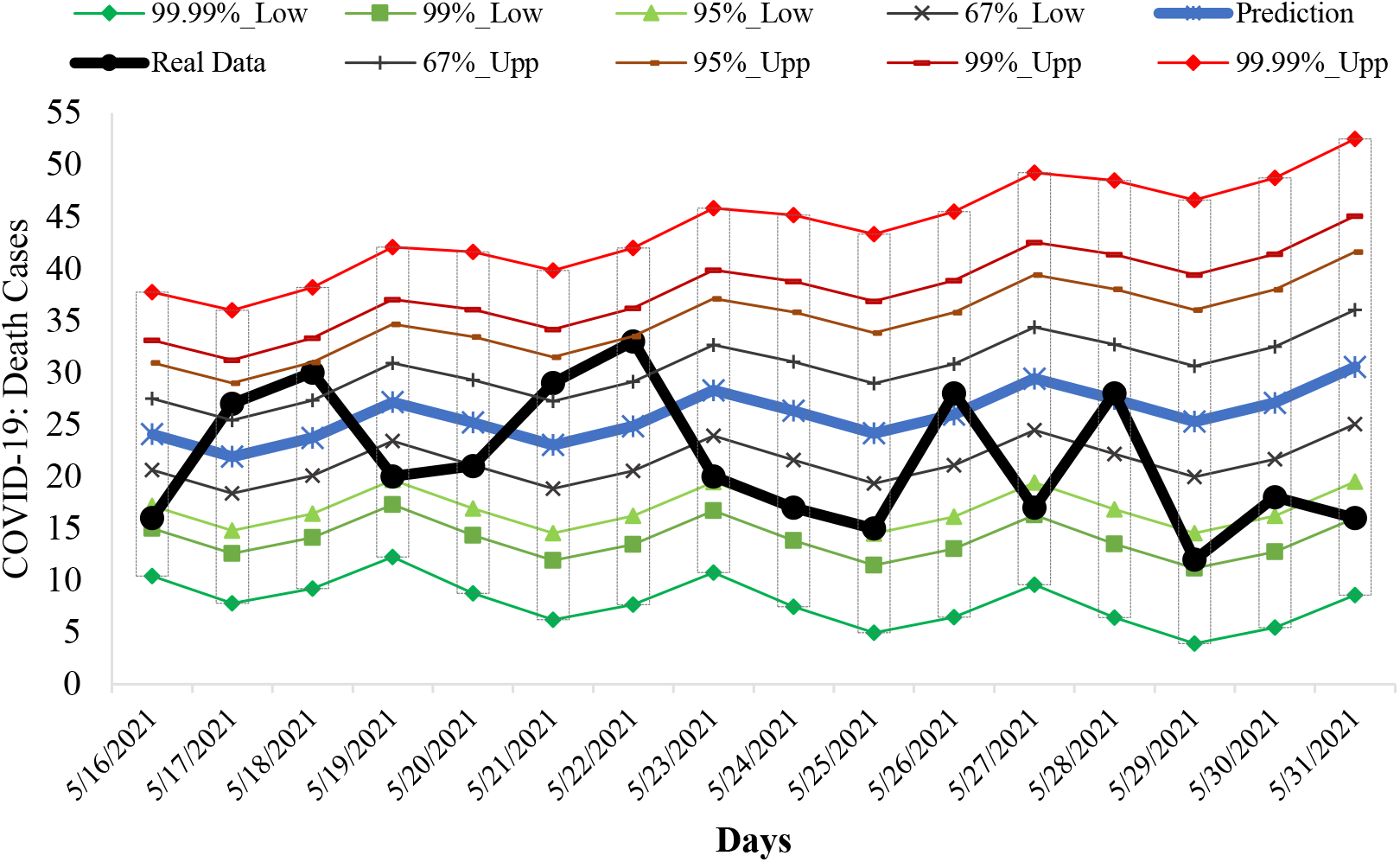
Example of Verification Interval Prediction of COVID-19 Death Cases in Georgia from 16.05.2021 to 31.25.2021.

**Fig. 23.**
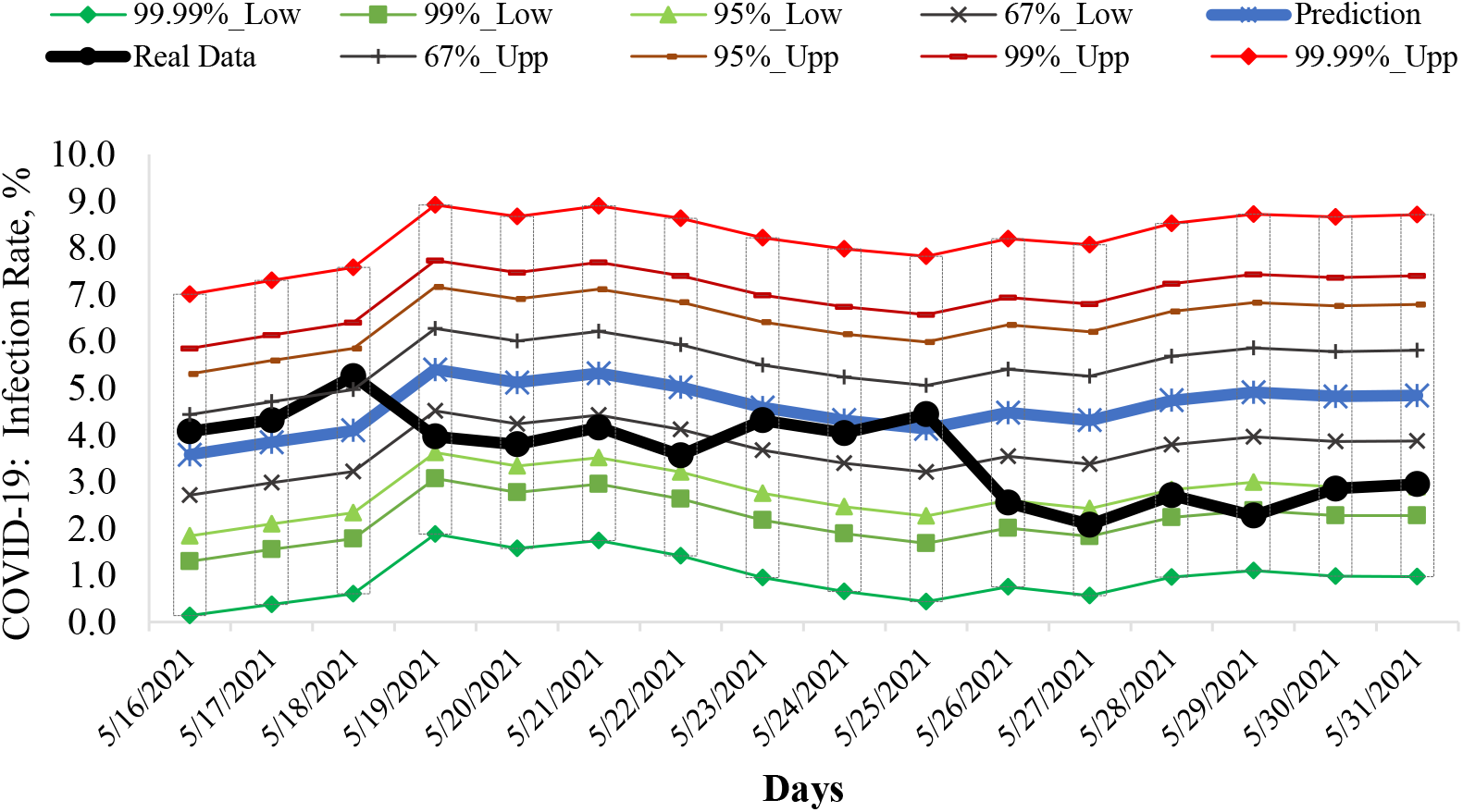
Example of Verification Interval Prediction of COVID-19 Infection Rate in Georgia from 16.05.2021 to 31.05.2021.

**Table 8.**
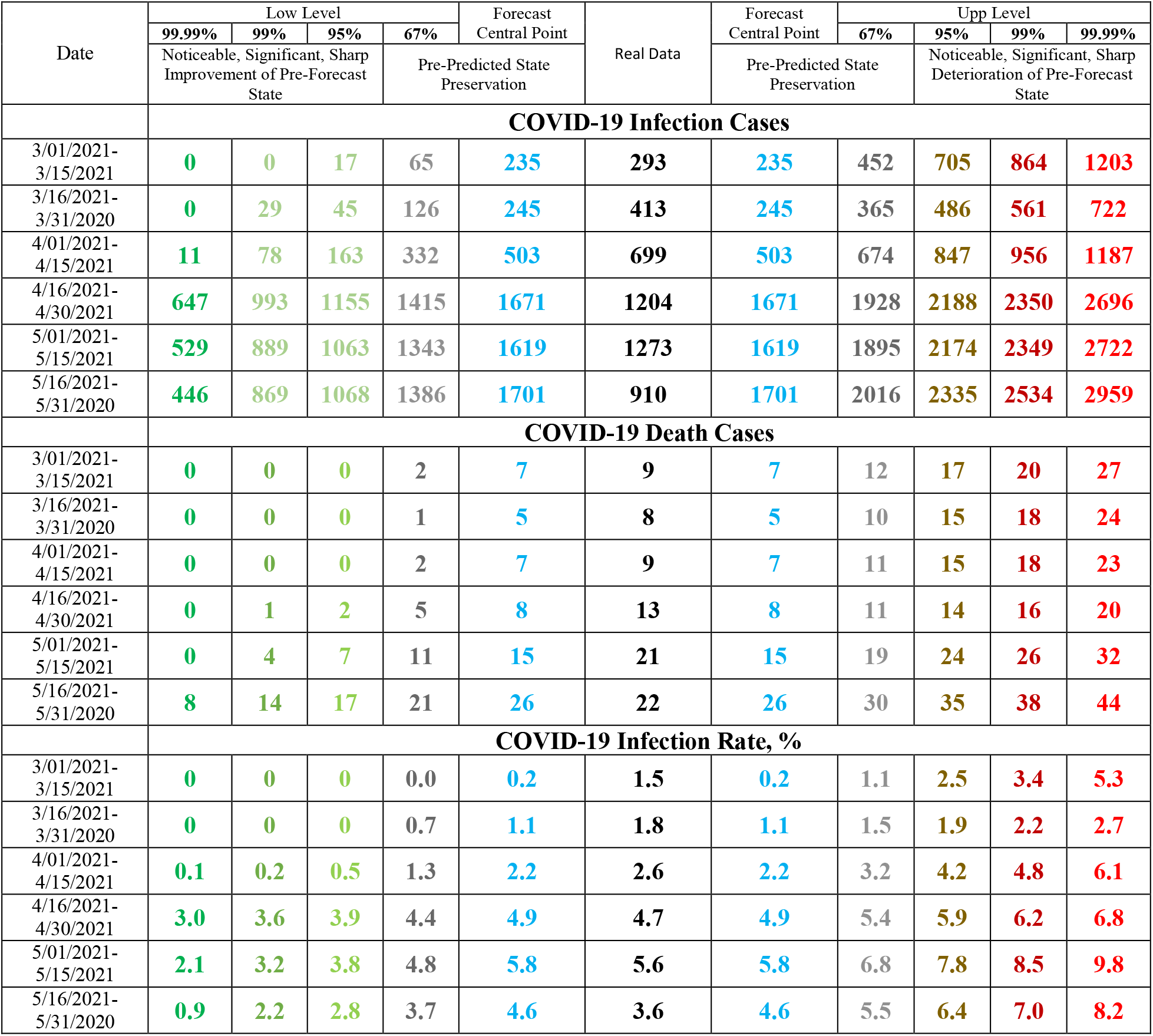
Verification of decade and two-week interval prediction of COVID-19 confirmed infection cases, deaths cases and infection rate in Georgia from 01.03.2021 to 31.05.2021.

Comparison of real and calculated predictions data of C, D and I shown that two-week daily and mean two-week real values of C, D and I practically falls into the 67% - 99.99% confidence interval of these predicted values for the specified time periods (Fig. 20-23, data from https://www.facebook.com/Avtandil1948/, Table 8).

For all forecast periods (6 forecasting cases), the stability of real time series of observations of these parameters (period for calculating the forecast + forecast period) remained (Table 9).

**Table 9.**
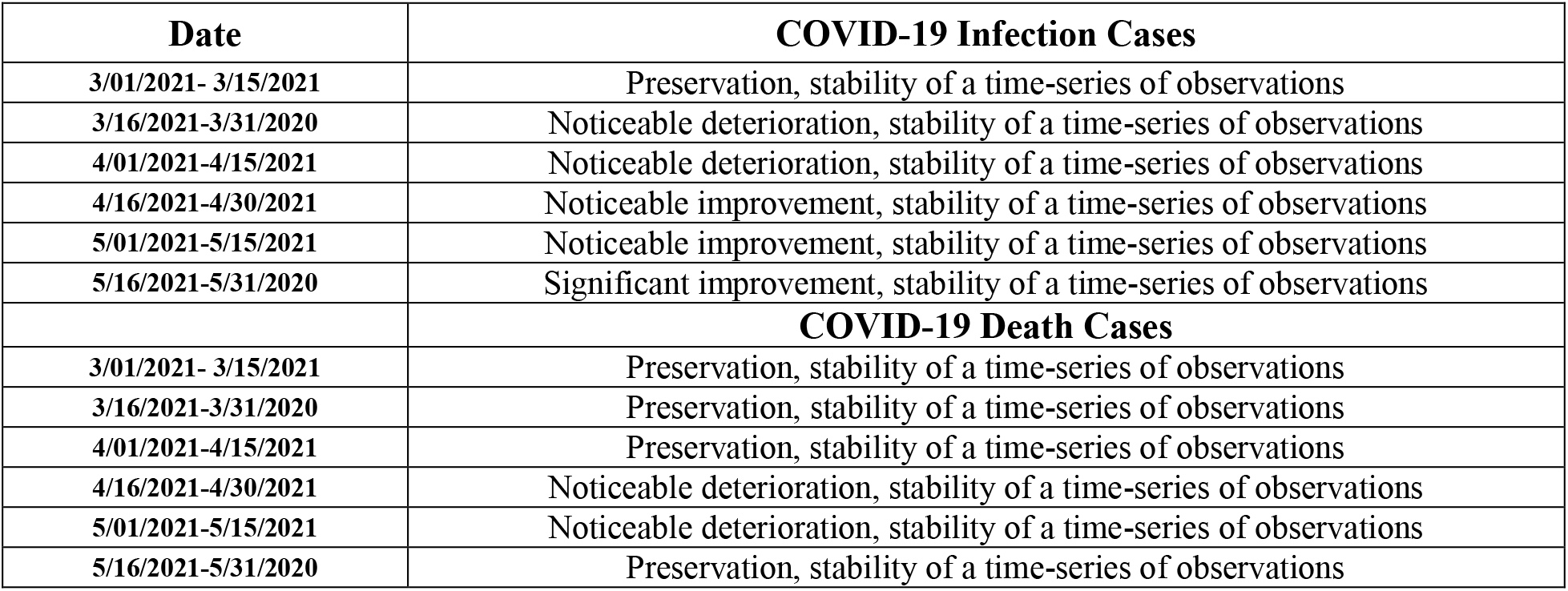
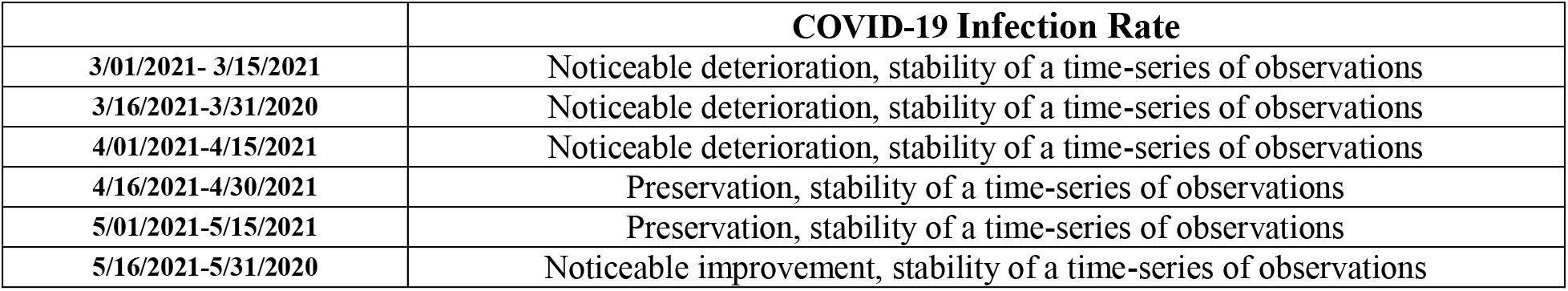
Change in the forecast state of C, D and I in relation to the pre-predicted one according to Table 1 scale [8].

Thus, the daily two-week and mean two-week forecasted values of C, D and I quite adequately describe the temporal changes in their real values. This was also confirmed by last verification of interval prediction of COVID-19 confirmed infection cases, deaths cases and infection rate in Georgia from 01.06.2021 to 15.06.2021 [https://www.facebook.com/Avtandil1948/].

## Conclusion

In the future, it is planned to continue regular similar studies for Georgia in comparison with neighboring and other countries, in particular, checking the temporal representativeness of short-term interval statistical forecasting of the new coronavirus infection COVID-19, taking into account data on the periodicity, clarification of connections between various parameters of this infection, etc.

## Data Availability

https://stopcov.ge

https://www.facebook.com/Avtandil1948/

## References

1. World Health Organization. (2020). Coronavirus Disease 2019 (COVID-19). Situation report. 67.

2. Öztoprak F, Javed A. (2020). Case fatality rate estimation of COVID-19 for European countries: Turkey’s current scenario amidst a global pandemic. Comparison of outbreaks with European countries. EJMO. 4(2): 149–159, DOI: 10.14744/ejmo.2020.60998.

3. Zemtsov SP, Baburin VL. (2020). Risks of morbidity and mortality during the COVID-19 pandemic in Russian regions. Population and economics. 4(2): 158-181. Available from: https://doi.org/10.3897/popecon.4.e54055

4. Meister S, Eradze I, Grigoryan A, Samadov B. The COVID-19 pandemic in the South Caucasus. ETH Zurich Research Collection. Available from: https://doi.org/10.3929/ethz-b-000415805

5. Covid-19 in Georgia. (2020). National Center for Disease Control & Public Health. 4 review. 64 p. (in Georgian).

6. Amiranashvili A.G, Khazaradze K.R, Japaridze N.D. (2020). Twenty weeks of the pandemic of coronavirus Covid-19 in Georgia and neighboring countries (Armenia, Azerbaijan, Turkey, Russia). Preliminary comparative statistical data analysis. Int. Sc. Conf. "Modern Problems of Ecology”, Proc., ISSN 1512-1976, v. 7, Tbilisi-Telavi, Georgia, 26-28 September, 2020, pp. 364–370.

7. Amiranashvili A.G., Khazaradze K.R., Japaridze N.D. (2020). Analysis of twenty-week time-series of confirmed cases of New Coronavirus COVID-19 and their simple short-term prediction for Georgia and neighboring countries (Armenia, Azerbaijan, Turkey, Russia) in amid of a global pandemic. medRxiv preprint doi: https://doi.org/10.1101/2020.09.09.20191494, p13 p. Europe PMC, https://europepmc.org/article/ppr/ppr213467

8. Amiranashvili A.G., Khazaradze K.R., Japaridze N.D. (2021). The Statistical Analysis of Daily Data Associated with Different Parameters of the New Coronavirus COVID-19 Pandemic in Georgia and their Short-Term Interval Prediction from September 2020 to February 2021. medRxiv preprint doi: https://doi.org/10.1101/2021.04.01.21254448, p18 p.

9. Padmabati Gahan, Monalisha Pattnaik, Agnibrata Nayak, Monee Kieran Roul. (2021). Prediction of COVID-19 pandemic of top ten countries in the world establishing a hybrid AARNN LTM model. 24 p. medRxiv preprint doi: https://doi.org/10.1101/2020.12.31.20249105

10. Yi Zhang, Sanjiv Kapoor. (2021). Hidden parameters impacting resurgence of SARS-CoV-2 pandemic. 20 p. medRxiv preprint doi: https://doi.org/10.1101/2021.01.15.20248217

11. Hazra L.K., Pujari B.S., Shekatkar S.M., Mozaffer F., Sinha S., Guttal V., Chaudhuri P., Menon G.I. (2021). The INDSCI-SIM Model for COVID-19 in India. 43 p. medRxiv preprint doi: https://doi.org/10.1101/2021.06.02.21258203

12. Abolmaali S., Shirzaei S. (2021). Forecasting COVID-19 Number of Cases by Implementing ARIMA and SARIMA with Grid Search in United States. 10 p. medRxiv preprint doi: https://doi.org/10.1101/2021.05.29.21258041

13. Yagoub R., Eledum H. (2021). Modeling of the COVID-19 Cases in Gulf Cooperation Council (GCC) Countries Using ARIMA and MA-ARIMA Models. 20 p. medRxiv preprint doi: https://doi.org/10.1101/2021.05.27.21257916

14. Roy S., Bhunia G. S., Shit P. K. (2021). Spatial Prediction of COVID-19 Epidemic Using ARIMA Techniques in India. Modeling Earth Systems and Environment, 7(2), pp. 1385–1391, doi.org/10.1007/s40808-020-00890-y.

15. Oladunni T., Denis M., Ososanya E., Adesina J. (2021). A Time Series Analysis and Forecast of COVID-19 Healthcare Disparity. 13 p. medRxiv preprint doi: https://doi.org/10.1101/2021.05.13.21257189

16. Förster E., Rönz B. (1979). Methoden der korrelations - und regressionsanalyse. – Ein Leitfaden für Ökonomen. Verlag Die Wirtshaft Berlin. 324.

17. Kendall MG. (1981). Time-series. Moscow, 200, (in Russian).

